# Loss of contractile arteriolar smooth muscle cells and their *GLP1R* expression in human diabetic kidney disease

**DOI:** 10.64898/2026.11.18.26368399

**Authors:** Shaun G. Hong, Myeongseop Kim, Donggun Kim, Hyunah Ku, Ara Ko, Jae-Ik Oh, Jinsun Lee, Min Woo Kang, Youmin Shin, Jeong Ho Joo, Jeong Seok Lee, Hajeong Lee, Seung Seok Han, Yaerim Kim, Jinkyung Kwon, Jeonghwan Lee, Jung Pyo Lee, Ji In Park, Jung Tak Park, Jung Nam An, Kyung-Chul Moon, Hyun Je Kim, Young-Gon Kim, Sehoon Park, Dong Ki Kim, KORNERSTONE investigators

## Abstract

Glucagon-like peptide-1 (GLP-1) receptor agonists slow kidney disease progression in type 2 diabetes, and in the human kidney the receptor is localized mainly to arteriolar smooth muscle. Whether these cells and their receptor are preserved in diabetic kidney disease (DKD) is unknown. We combined single-cell-level spatial transcriptomics of 5,101 genes with registered periodic acid–Schiff histology in kidney biopsies from 28 people with DKD, 102 with other glomerular diseases and 24 living donors, and measured 2,531 arterioles and small arteries. Arteriolar walls in DKD contained about 20% fewer smooth muscle cells than those in other glomerular diseases, and the loss fell on cells without matrix gene expression. Transcripts of *GLP1R*, which encodes the receptor, were detected in 0.48% of the remaining smooth muscle cells in DKD, compared with 1.21% in other glomerular diseases and 1.75% in donors. The glutamate receptor gene *GRIA2*and the smooth muscle regulator *PRDM6* were also lower, whereas contractile genes were unchanged, a pattern not seen in the other diseases. In an independent single-nucleus atlas, smooth muscle *GLP1R* was lower in DKD than in reference kidneys, and smooth muscle content fell with arteriolar hyalinosis grade. In human DKD, arterioles lose contractile smooth muscle cells, and fewer of the remaining cells carry transcripts for the receptor targeted by GLP-1 receptor agonists at the disease stage in which these drugs are used.

**One Sentence Summary:** In human diabetic kidney disease, kidney arterioles lose contractile smooth muscle cells, and fewer of the remaining cells express the GLP-1 receptor.

## 1 Introduction

Diabetic kidney disease (DKD) is a leading cause of kidney failure, with injury to preglomerular vessels a defining feature. Arteriolar hyalinosis is graded in the pathological classification of diabetic nephropathy [1], is associated with cardiovascular events [2] and increases with age across kidney diseases [3]. Smooth muscle is thought to be replaced by hyaline material [1], but morphology alone cannot distinguish fewer cells from more matrix between preserved cells. Molecular correlates of the lesion are only beginning to be defined [4].

The same vessels have become relevant to treatment. In the FLOW trial, the glucagon-like peptide-1 (GLP-1) receptor agonist semaglutide slowed kidney disease progression in people with type 2 diabetes and chronic kidney disease [5], extending earlier kidney signals seen with liraglutide [6] and dulaglutide [7]. In the human kidney, an extensively validated monoclonal antibody localized the GLP-1 receptor to the smooth muscle of preglomerular arteries and arterioles rather than to tubular epithelium [8], and this localization has remained the reference since earlier antibodies proved unreliable [9]. In experimental models, receptor activation on vascular smooth muscle reduces afferent arteriolar autoregulation and increases kidney blood flow [10], induces renin along the vascular tree [11] and lowers blood pressure through the smooth muscle receptor [12]. In the REMODEL trial, semaglutide reduced the intimal area of the most diseased arterioles after one year [13].

Arteriolar smooth muscle is therefore both a principal site of injury in DKD and the main site of the GLP-1 receptor in the kidney, yet whether these cells and their receptor are preserved in human DKD is unknown.

In this study, we combined single-cell-level spatial transcriptomics with registered periodic acid–Schiff (PAS) histology in kidney biopsies from people with DKD, people with other glomerular diseases and living donors. We counted smooth muscle cells in the walls of arterioles and small arteries, defined which smooth muscle cells were lost, measured *GLP1R* and other mural genes in the cells that remained, and compared DKD with other glomerular diseases to separate changes associated with diabetes from those shared with glomerular injury.

## 2 Results

### 2.1 Cohort and arterioles

The spatial cohort contained 5,195,294 cells from 154 participants: 28 with DKD, 102 with other glomerular diseases (60 with IgA nephropathy [IgAN], 25 with minimal change disease [MCD], 13 with membranous nephropathy [MN] and four with focal segmental glomerulosclerosis [FSGS]) and 24 living kidney donors (Table 1; Fig. 1a). All living donors had an eGFR above 60 mL/min/1.73 m2 and a urine protein-to-creatinine ratio below 0.15 g/g, none had diabetes, and 3 of 24 had hypertension (Table 1). Participants with DKD had lower estimated glomerular filtration rate (eGFR) than those with other glomerular diseases (median 54.1 versus 93.2 to 103.5 mL/min/1.73 m^2^ in IgAN, MCD and MN) and more often had hypertension (18 of 24 versus 27 of 82); 19 of the 28 had an eGFR between 25 and 75 mL/min/1.73 m^2^. The morpho-transcriptomic definition identified 2,531 arterioles and small arteries in 144 participants, including 417 from all 28 participants with DKD (fig. S1). Registered PAS walls coincided with *ACTA2* -expressing mural cells and an endothelial lining in the spatial data (Fig. 1b).

**Fig. 1.**
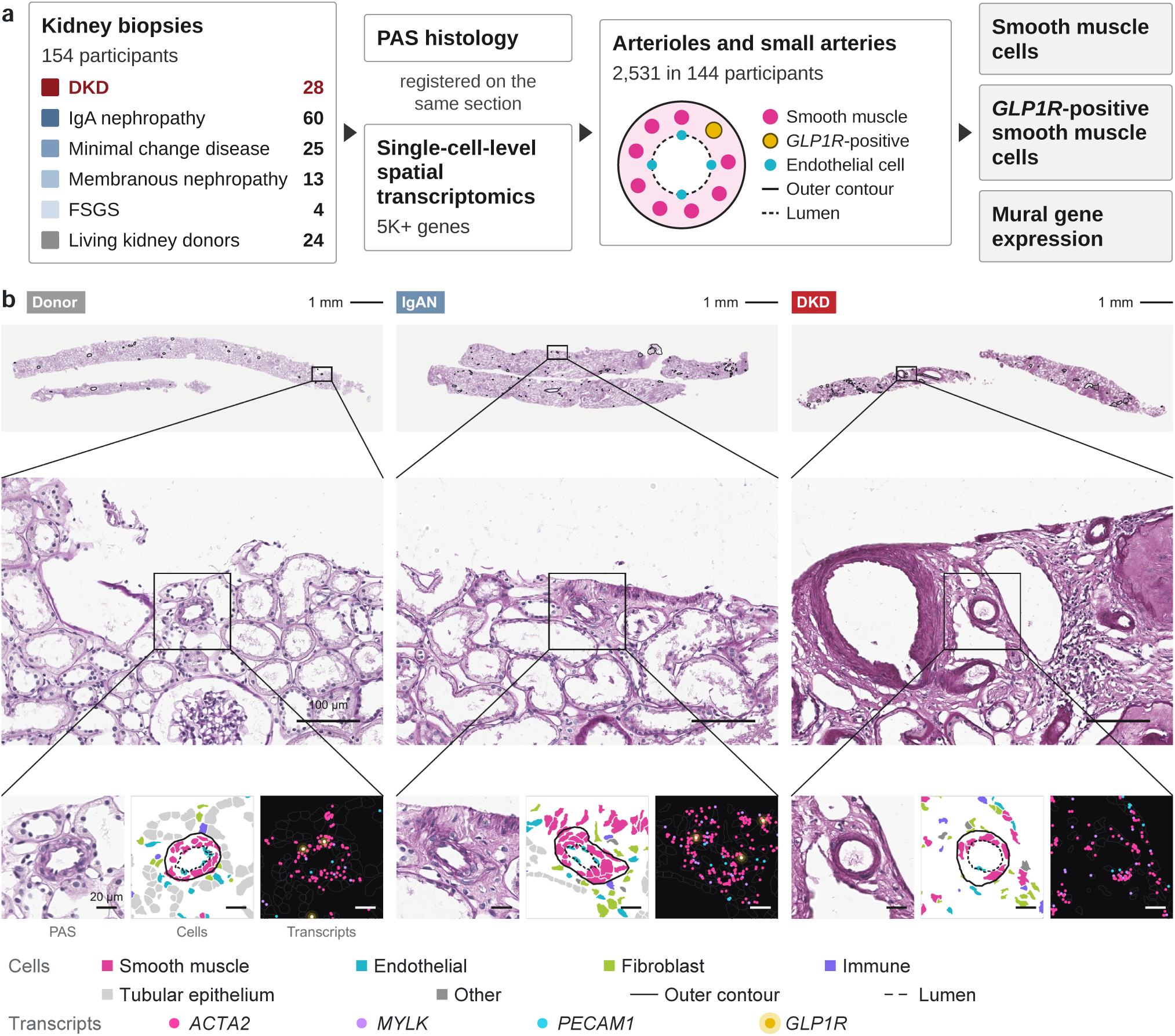
Arterioles in human kidney biopsies. (a) Study overview. One biopsy section per participant from 28 participants with diabetic kidney disease (DKD), 24 living donors, 60 with IgA nephropathy (IgAN), 25 with minimal change disease (MCD), 13 with membranous nephropathy (MN) and four with focal segmental glomerulosclerosis (FSGS) underwent single-cell-level spatial transcriptomics with registered PAS histology. A morpho-transcriptomic definition, combining PAS morphology with the transcriptomic identity of wall cells, identified 2,531 arterioles and small arteries in 144 participants; smooth muscle cells in the wall, *GLP1R*-positive smooth muscle cells and mural gene expression were measured. (b) Registered PAS histology, cell identities and transcript calls in representative donor, IgAN and DKD sections at three magnifications. Thin outlines mark all segmented vascular profiles in the section. Solid outlines mark the outer contour and dashed outlines the lumen; cell colors identify smooth muscle, endothelial, fibroblast, immune, tubular epithelial and other cells; transcript colors distinguish *ACTA2*, *MYLK*, *PECAM1* and *GLP1R*. Scale bars are shown within each field.

**Table 1.** Characteristics of the participants.

| Characteristic | DKD | Living donors | IgAN | MCD | MN | FSGS |
|---|---|---|---|---|---|---|
| Participants, *n* | 28 | 24 | 60 | 25 | 13 | 4 |
| Age, years | 51.3 (38.4–59.8) | 55.0 (50.0–58.2) | 39.6 (33.8–50.1) | 41.8 (31.4–57.0) | 59.7 (53.0–65.3) | 31.9 (20.3–44.9) |
| Female, *n* (%) | 8 (28.6) | 17 (70.8) | 30 (50.0) | 9 (36.0) | 4 (30.8) | 4 (100.0) |
| eGFR, mL/min/1.73 m<sup>2</sup> | 54.1 (39.6–84.2) | 102.9 (98.8–106.0) | 93.2 (79.2–111.7) | 103.5 (81.2–115.5) | 102.9 (97.2–106.3) | 78.9 (48.1–114.0) |
| UPCR, g/g | 3.99 (3.04–8.21); *n* = 27 | 0.05 (0.03–0.06) | 0.86 (0.49–1.19) | 8.55 (5.80–11.63) | 4.78 (4.07–7.07) | 4.35 (1.73–8.10) |
| Hypertension, *n*/*N* (%) | 18/24 (75.0) | 3/24 (12.5) | 21/60 (35.0) | 2/12 (16.7) | 4/10 (40.0) | — |
| Diabetes, *n*/*N* (%) | 24/24 (100.0) | 0/24 (0.0) | 0/60 (0.0) | 0/12 (0.0) | 2/10 (20.0) | — |
| Systolic blood pressure, mmHg | 123.0 (110.5–134.5); *n* = 23 | — | 113.0 (104.8–123.0) | 119.0 (113.0–123.0); *n* = 11 | 118.5 (107.5–124.5); *n* = 10 | — |
| Body mass index, kg/m<sup>2</sup> | 23.8 (22.0–26.8); *n* = 23 | 24.1 (22.2–25.5) | 23.6 (21.8–25.6) | 24.7 (23.3–25.9); *n* = 11 | 25.7 (22.2–26.8); *n* = 10 | — |
| Pre-biopsy RAS inhibitor, *n*/*N* (%) | 15/23 (65.2) | — | 26/60 (43.3) | 4/11 (36.4) | 6/10 (60.0) | — |
| Arterioles per participant | 11 (7–24) | 17 (11–21) | 17 (10–32) | 10 (5–16) | 10 (7–15) | 16 (14–18) |

Values are median (interquartile range) or counts and percentages; observed sample sizes are given where incomplete. Donor characteristics come from donor records and the pre-donation evaluation; donor eGFR was calculated from pre-donation creatinine with the 2021 CKD-EPI equation, and donor UPCR was measured a median of 98 days before donation. Urine dipstick albumin within 22 days before donation was negative in 22 donors and trace in 2. Hypertension and diabetes are shown as number with the condition / number with a recorded history (percentage). Arteriole counts per participant describe the 144 participants with eligible profiles. One primary section represents each participant. DKD, diabetic kidney disease; IgAN, IgA nephropathy; MCD, minimal change disease; MN, membranous nephropathy; FSGS, focal segmental glomerulosclerosis; eGFR, estimated glomerular filtration rate; UPCR, urine protein-to-creatinine ratio; RAS, renin–angiotensin system. A dash denotes an unavailable measurement; FSGS hypertension and diabetes histories and donor systolic blood pressure and medication use were not recorded. RAS inhibitor use is given among participants with pre-biopsy medication records (DKD 23 of 28, IgAN 60 of 60, MCD 11 of 25, MN 10 of 13).

### 2.2 Fewer contractile smooth muscle cells in diabetic arteriolar walls

Smooth muscle density was lower in DKD than in other glomerular diseases (−1,941 cells/mm^2^ of wall; 95% confidence interval [CI], −2,544 to −1,338), about 20% of the comparator mean (Holm-adjusted *P* < 0.001; Fig. 2, a to c; Table 2). The difference was −1,413 cells/mm^2^ (−2,022 to −804) after age, sex and batch adjustment, and the deficit was also seen per 100 µm of outer perimeter (Table 2). The deficit was present against donors and against other diagnoses on the same slides (table S1).

**Fig. 2.**
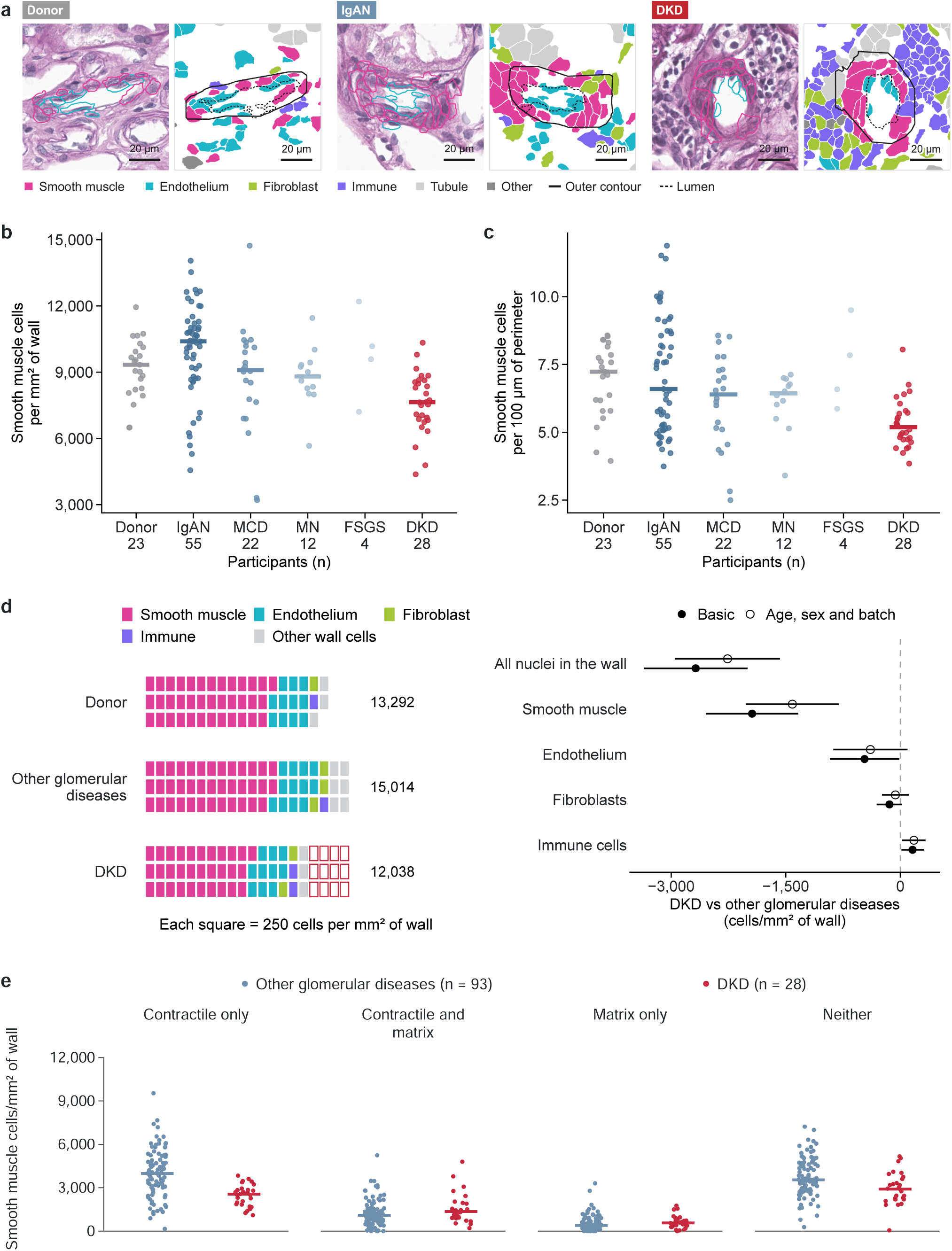
Fewer contractile smooth muscle cells in diabetic arteriolar walls. (a) PAS and corresponding cell segmentation in arterioles from a donor, IgAN and DKD, selected within ±10 µm of the median DKD outer diameter in the largest caliber and near each group’s median smooth muscle density. Scale bars, 20 µm. (b and c) Participant mean smooth muscle density per wall area (b) and count per 100 µm of outer perimeter (c). Points are participants and horizontal bars medians; FSGS is shown as points alone. Numbers beneath the diagnoses are participants with eligible arterioles. (d) Wall-cell composition and differences by cell class. Each filled square represents 250 cells/mm^2^ of wall in the group mean; outlined squares show the DKD deficit relative to the mean of other glomerular diseases. Filled points are basic estimates and open points add age, sex and batch, with 95% CIs. (e) Wall density of arteriolar smooth muscle cells with at least one nuclear transcript, by transcript status: contractile (≥1 transcript of the contractile-score genes) and matrix (≥1 transcript of *COL1A1*, *COL1A2*, *FN1* or *TNC*). Points are participants and bars medians. Table 2 and table S1 report cellularity and matrix-fraction models; source data for (e) are in data file S1.

**Table 2.** Arteriolar wall cells and *GLP1R* in DKD.

| Measurement | Basic effect (95% CI) | *q* | Adjusted effect (95% CI) | *q* |
|---|---|---|---|---|
| Smooth muscle cells per mm<sup>2</sup> of wall | −1,941 (−2,544 to −1,338) | 9.99 × 10<sup>−10</sup> | −1,413 (−2,022 to −804) | 1.25 × 10<sup>−5</sup> |
| Smooth muscle cells per 100 µm of perimeter | −1.35 (−1.98 to −0.72) | 3.74 × 10<sup>−5</sup> | −0.92 (−1.57 to −0.26) | 0.008 |
| All nuclei in the wall per mm<sup>2</sup> | −2,679 (−3,357 to −2,001) | 6.66 × 10<sup>−14</sup> | −2,261 (−2,948 to −1,575) | 7.49 × 10<sup>−10</sup> |
| *ACTA2*-positive cells per mm<sup>2</sup> | −2,058 (−3,017 to −1,100) | 3.74 × 10<sup>−5</sup> | −1,091 (−2,041 to −141) | 0.028 |
| *GLP1R*-positive smooth muscle cells (rate ratio) | 0.33 (0.17 to 0.66) | 0.010 | 0.36 (0.18 to 0.69) | 0.011 |
| *GLP1R*-positive cells per arteriole (rate ratio) | 0.34 (0.14 to 0.81) | 0.023 | 0.34 (0.13 to 0.88) | 0.038 |
| Arteriolar smooth muscle *GLP1R* expression (ratio) | 0.54 (0.39 to 0.74) | 0.007 | 0.55 (0.40 to 0.77) | 0.008 |

DKD versus other glomerular diseases. Differences are absolute for cellularity and ratios for detection, positive cells per arteriole and expression. Basic estimates include diagnosis and, for profile measurements, caliber; adjusted estimates add age, sex and batch. The comparator is the mean of the other glomerular diseases, weighted by the number of participants. All nuclei and *ACTA2*-positive cells are counted irrespective of lineage annotation. *q* values are Benjamini–Hochberg values across seven cellularity tests within each model (the four cellularity rows, non-smooth-muscle wall cells (Fig. 2d) and the two donor rows of table S1) and across the *GLP1R* analyses (table S4). There were 2,531 arterioles in 144 participants and 29,594 arteriolar smooth muscle cells. Donor comparisons are in tables S1 and S4.

The lower density reflected fewer cells rather than a change in cell identity. All nucleated wall cells were fewer by 2,679 cells/mm^2^ (−3,357 to −2,001) and cells identified by *ACTA2* transcripts alone by 2,058 (−3,017 to −1,100), whereas non-smooth-muscle cells were not more numerous (−479; −1,042 to 83; Fig. 2d).

Matrix gene expression separated the cells that were lost from those that remained. Among cells with at least one nuclear transcript, 27.3% of DKD arteriolar smooth muscle cells carried at least one *COL1A1*, *COL1A2*, *FN1* or *TNC* transcript, compared with 20.2% in other glomerular diseases (ratio, 1.53; 1.26 to 1.85), whereas the proportion carrying contractile transcripts did not differ (0.93; 0.83 to 1.04). Counted per wall area, the deficit fell on cells with contractile transcripts only (−1,476 cells/mm^2^; −2,047 to −904) and on cells with neither (−773; −1,288 to −258), whereas cells carrying matrix transcripts were not fewer (Fig. 2e). Accordingly, the remaining smooth muscle cells had a higher matrix score (0.61; 0.32 to 0.89) with an unchanged contractile score (−0.04; −0.28 to 0.21). Matrix scores did not differ in fibroblasts (0.17; −0.45 to 0.79) or endothelium (−0.06; −0.27 to 0.16) of the same walls (table S2).

The loss tracked the wall lesion. Within arterioles, each 10 percentage points of PAS-dense acellular wall was associated with 793 fewer smooth muscle cells/mm^2^ (−934 to −653), including within DKD (−579; −860 to −297; table S1). The DKD deficit persisted after adjustment for eGFR and proteinuria or for hypertension (table S1). Adjustment for recorded arteriolar hyalinosis attenuated the deficit (−1,416; −2,531 to −302), and its interval included zero once age, sex and batch were added (−855; −1,974 to 265). Thick-walled profiles with fewer than three smooth muscle cells, which lay outside the primary set, were more frequent in DKD, and including them enlarged the deficit (−2,440 cells/mm^2^; table S1).

### 2.3 Fewer *GLP1R*-positive smooth muscle cells in diabetic arteriolar walls

*GLP1R* detection was higher in arteriolar smooth muscle than in mural cells outside the outlined arterioles (356 of 29,229 versus 1,010 of 225,415 cells), and *GLP1R* transcripts were rarely detected in renin-associated cells (7 of 7,122; Fig. 3b). In DKD, 0.48% of arteriolar smooth muscle cells (20 of 4,145) were *GLP1R*-positive, compared with 1.21% (247 of 20,371) in other glomerular diseases and 1.75% (89 of 5,078) in donors (Fig. 3, a and c; table S3). The ratio of *GLP1R* detection rates relative to other glomerular diseases was 0.33 (95% CI, 0.17 to 0.66; Holm-adjusted *P* = 0.002) and 0.36 (0.15 to 0.85) after adjustment for age, sex, batch, eGFR and hypertension (Table 2 and table S4). The reduction was consistent across participant-level and background-aware models and within batch and caliber subsets (table S4).

**Fig. 3.**
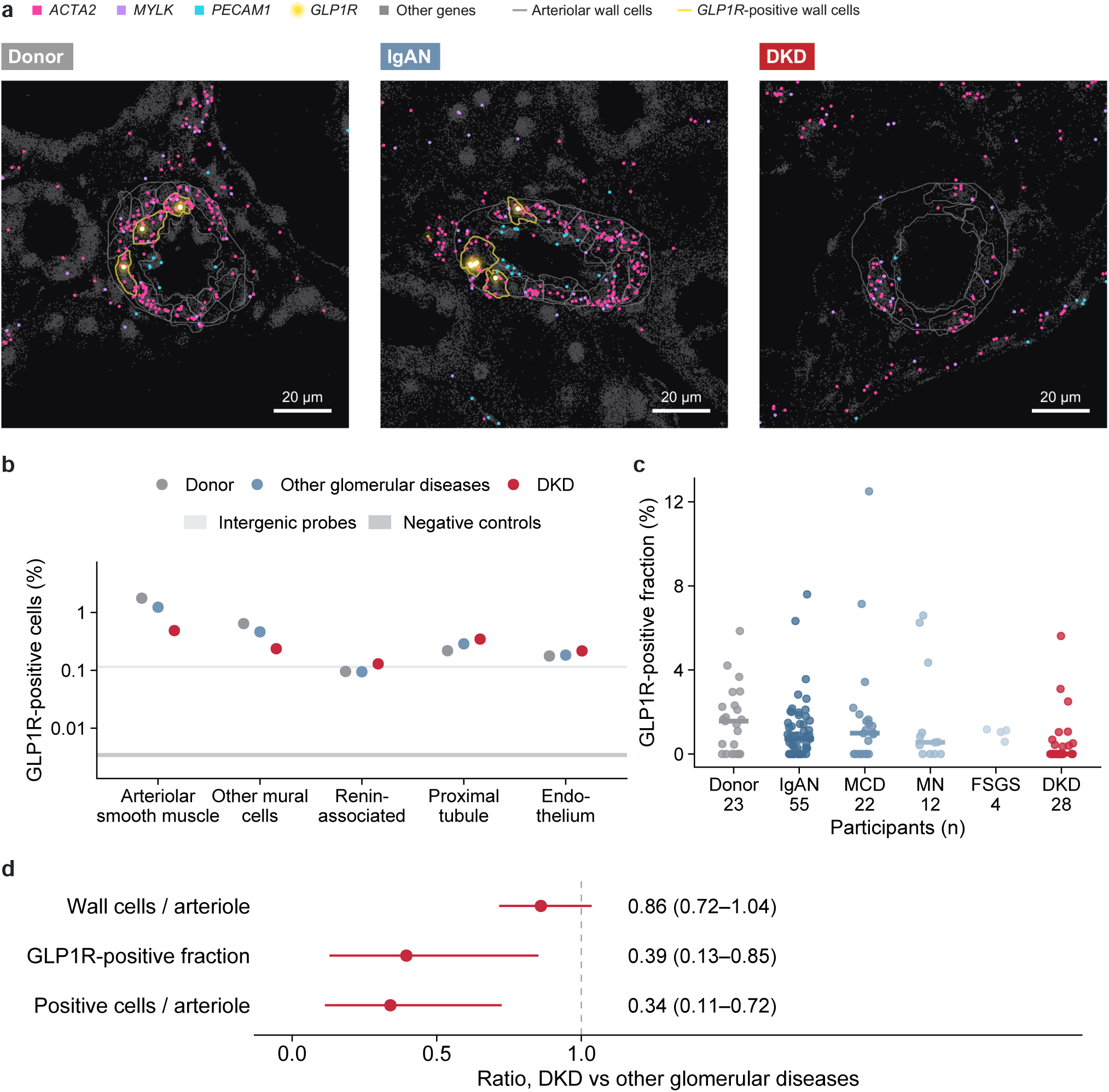
Fewer *GLP1R*-positive smooth muscle cells in diabetic arteriolar walls. (a) Transcript maps of donor, IgAN and DKD arterioles in equally sized 120-µm fields. Yellow outlines identify arteriolar smooth muscle cells with at least one nuclear *GLP1R* transcript; gray outlines mark the remaining arteriolar smooth muscle cells. Scale bars, 20 µm. (b) Pooled nuclear *GLP1R*-positive fractions in arteriolar smooth muscle (356 of 29,229 cells), mural cells outside the outlined arterioles (1,010 of 225,415), renin-associated cells (7 of 7,122), proximal tubule and endothelium, by diagnosis group. Bands show the range of false-positive rates expected from intergenic and negative-control probes. The ordinate is logarithmic. (c) Participant arteriolar smooth muscle *GLP1R* positivity across six diagnoses. Points are participants and bars medians; FSGS is shown as points alone. (d) Decomposition of *GLP1R*-positive cells per arteriole into cells per arteriole and the *GLP1R*-positive fraction. Points are pooled ratios with participant-bootstrap 95% CIs; these intervals differ slightly from the model-based intervals in Table 2. Lower positivity contributed 86% and fewer smooth muscle cells 14% of the difference.

Wall positivity exceeded the false-positive rate expected from intergenic probes 17.0-fold in donors, 11.2-fold in other glomerular diseases and 4.5-fold in DKD (95% CI, 1.5 to 8.7; fig. S2), whereas proximal-tubule, endothelial and intergenic-probe positivity did not differ between DKD and other glomerular diseases. *GLP1R*-positive cells per arteriole were 66% fewer in DKD (ratio, 0.34; 0.14 to 0.81); lower positivity per cell accounted for 86% of this difference and fewer cells per arteriole for 14% (Fig. 3d). Without the PAS outlines, mural-cell *GLP1R* expression was also lower in DKD (table S4). Relative to the 23 participants with hypertension but without diabetes, the adjusted ratio was 0.34 (0.13 to 0.91).

### 2.4 Selective changes in the remaining smooth muscle cells

Of the 118 mural genes, five differed between DKD and other glomerular diseases (Fig. 4a and Table 3). *GLP1R* (log_2_ fold change, −0.90; 95% CI, −1.36 to −0.44), *GRIA2* (−1.12; −1.54 to −0.69), *PRDM6* (−0.79; −1.22 to −0.36) and *REN* (−1.23; −1.98 to −0.48) were lower, whereas *P2RX1* remained at donor levels in DKD and was lower in the other glomerular diseases. The reductions in *GLP1R* and *GRIA2* persisted after adjustment for transcripts per cell and after excluding the two participants with other glomerular diseases who had diabetes (table S5). For *GLP1R*, *GRIA2* and *PRDM6*, IgAN, MCD, MN and FSGS showed no significant reduction relative to donors (Fig. 4b). Across the 48 panel vasoregulatory genes, including *AGTR1*, *EDNRA* and *CACNA1C*, only *GLP1R*, *GRIA2*, *PRDM6* and *REN* were lower in DKD than in other glomerular diseases, and *ACTA2* did not differ (fig. S3). Lower *REN* was not explained by renin–angiotensin system (RAS) inhibitor use before biopsy (−1.24; −1.94 to −0.54; table S5). After adjustment for transcript yield, extracellular-matrix structural constituents and transforming growth factor-*β* response genes were enriched in DKD smooth muscle, in line with its higher matrix score (Fig. 4c and table S2).

**Fig. 4.**
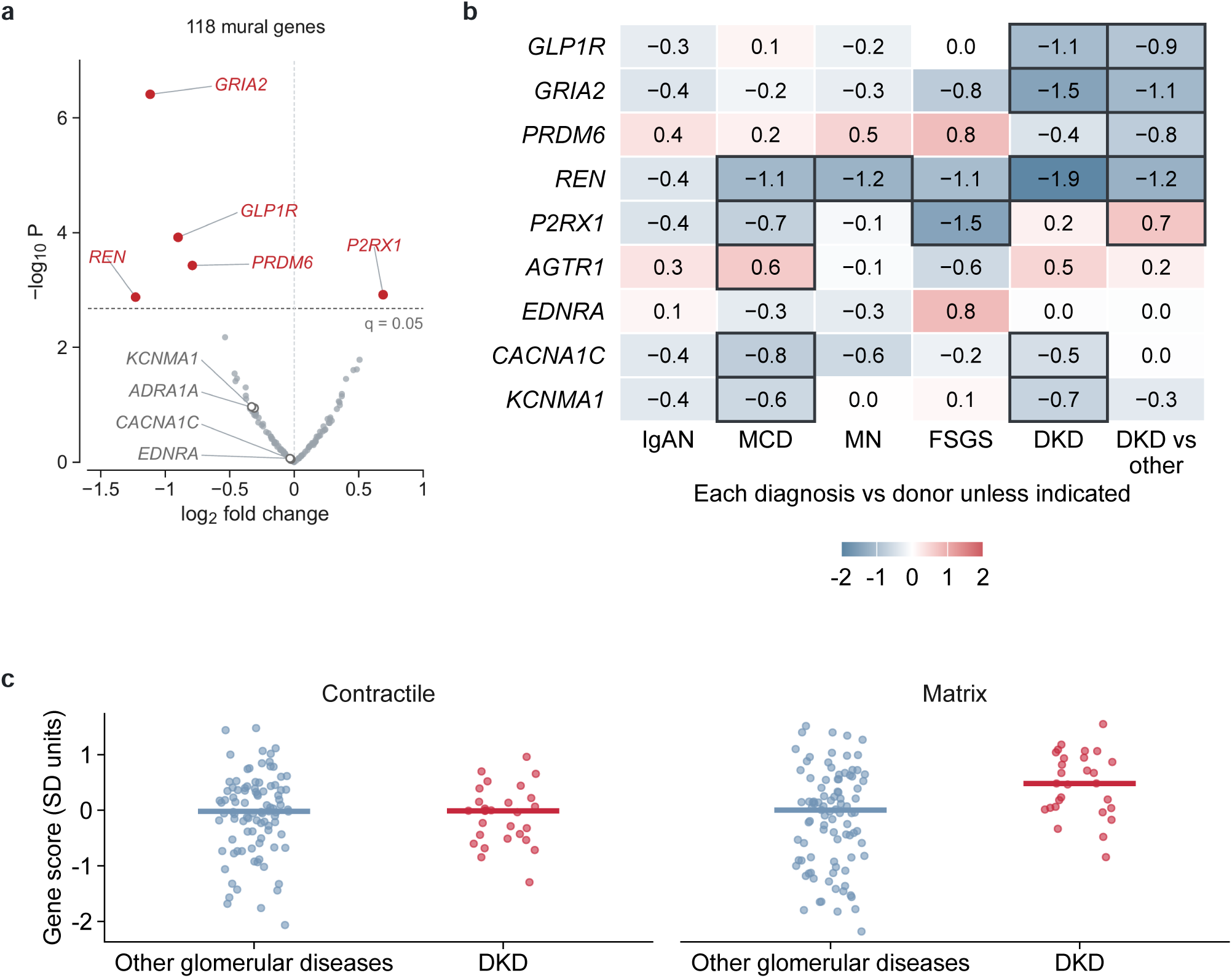
Selective changes in the remaining smooth muscle cells. (a) Differential expression among 118 mural genes, DKD versus other glomerular diseases, from participant pseudobulk. Red points pass false discovery rate control within the 118 genes; the five altered genes and contractile reference genes are labeled. (b) Diagnosis-specific effects for *GLP1R*, *GRIA2*, *PRDM6*, *REN*, *P2RX1* and four reference genes. Each disease is compared with donors, and the final column compares DKD with the participant-weighted mean of other glomerular diseases. Tiles show log_2_ fold change; borders indicate 95% CIs excluding zero. (c) Contractile and matrix gene scores in participants with other glomerular diseases and DKD. Points are participants and bars medians. Table 3 and tables S2 and S5 report gene effects, clinical sensitivities and gene-set enrichment; the complete receptor panel is in fig. S3.

**Table 3.** Selective mural-gene changes in diabetic arteriolar walls.

| Gene | Comparison | Basic log<sub>2</sub> FC (95% CI) | Adjusted log<sub>2</sub> FC (95% CI) |
|---|---|---|---|
| *GLP1R* | vs other glomerular diseases | −0.90 (−1.36 to −0.44) | −0.86 (−1.34 to −0.39) |
|  |  | *q* = 0.007 | *q* = 0.024 |
| *GRIA2* | vs other glomerular diseases | −1.12 (−1.54 to −0.69) | −1.08 (−1.54 to −0.63) |
|  |  | *q* = 4.58 × 10<sup>−5</sup> | *q* = 4.86 × 10<sup>−4</sup> |
| *PRDM6* | vs other glomerular diseases | −0.79 (−1.22 to −0.36) | −0.71 (−1.17 to −0.25) |
|  |  | *q* = 0.015 | *q* = 0.063 |
| *REN* | vs other glomerular diseases | −1.23 (−1.98 to −0.48) | −1.14 (−1.92 to −0.36) |
|  |  | *q* = 0.031 | *q* = 0.081 |
| *P2RX1* | vs other glomerular diseases | 0.69 (0.27 to 1.10) | 0.56 (0.12 to 1.00) |
|  |  | *q* = 0.031 | *q* = 0.199 |
| *ACTA2* | vs other glomerular diseases | −0.01 (−0.38 to 0.36) | 0.34 (−0.04 to 0.72) |
|  |  | *q* = 0.998 | *q* = 0.314 |

Participant wall-cell pseudobulk (nuclear transcripts, 144 participants); log<sub>2</sub> fold change. *q* values control the false discovery rate within the 118 mural-maximal genes for DKD versus other glomerular diseases, separately by model. *ACTA2* is shown as a reference. Adjusted models add age, sex and batch. Effects against donors, each individual diagnosis and 48 vasoregulatory receptor genes are in data file S1.

### 2.5 Independent cohort

In the Kidney Precision Medicine Project (KPMP) single-nucleus atlas, smooth muscle *GLP1R* was lower in DKD than in reference kidneys (log_2_ fold change, −1.18; 95% CI, −2.10 to −0.25; Fig. 5a), and *GLP1R*-positive smooth muscle nuclei numbered 5 of 622 versus 42 of 892 (Fig. 5b). *GRIA2* was also lower (−0.79; −1.54 to −0.04). *PRDM6*, by contrast, was higher in KPMP DKD than in reference kidneys (1.61; 1.00 to 2.21), and *REN* and *P2RX1* did not differ (Fig. 5a). Mural-gene effects against reference kidneys correlated between this study and KPMP (*ρ* = 0.65; fig. S4). The smooth muscle fraction fell with each grade of arteriolar hyalinosis (−0.47 log_2_ units per grade; −0.83 to −0.10; Fig. 5c).

**Fig. 5.**
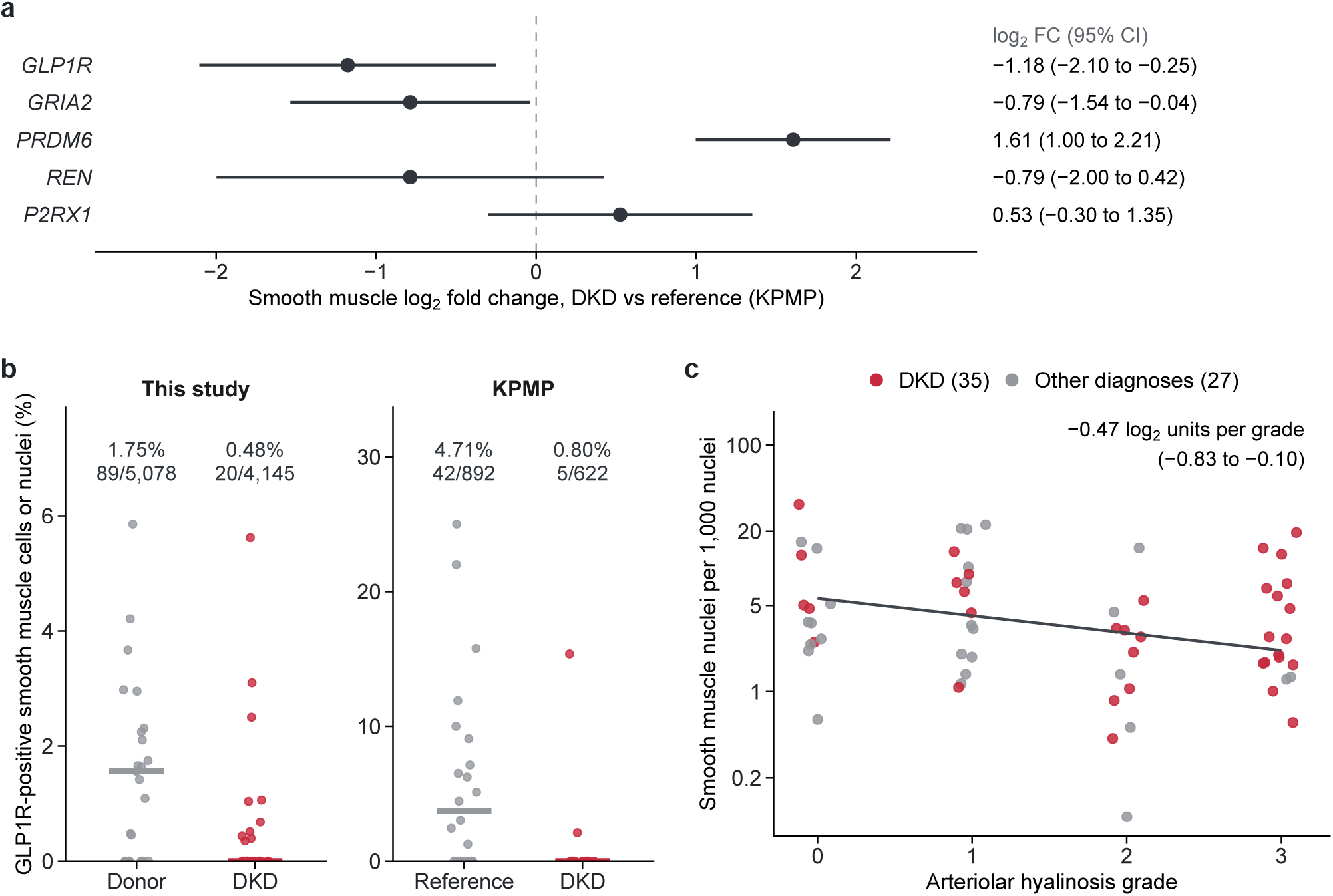
An independent cohort and arteriolar hyalinosis. (a) Vascular smooth muscle log_2_ fold changes for *GLP1R*, *GRIA2*, *PRDM6*, *REN* and *P2RX1* in DKD versus reference kidneys in the KPMP single-nucleus atlas, from participant pseudobulk with at least ten smooth muscle nuclei per participant; points are estimates with 95% CIs. *PRDM6* was higher in KPMP DKD, in the opposite direction to this study. (b) Participant *GLP1R*-positive smooth muscle fractions in this study and in KPMP. Points are participants and bars medians; pooled percentages and positive/total counts appear above the groups. (c) KPMP smooth muscle nuclei per 1,000 nuclei by arteriolar hyalinosis grade in 62 participants with available pathology; colors distinguish DKD from the other diagnoses. The line shows the age-, sex- and DKD-adjusted slope.

## 3 Discussion

In kidney biopsies from people with DKD, arterioles and small arteries contained fewer smooth muscle cells, and the cells that remained less often carried *GLP1R* transcripts. The cells that were lost were mainly contractile cells without matrix gene expression, so the remaining wall was weighted toward cells that express matrix genes. In the remaining cells, *GLP1R*, *GRIA2* and *PRDM6* were lower while contractile genes were preserved, a pattern that distinguished DKD from four other glomerular diseases sampled by the same biopsy route. Lower smooth muscle *GLP1R* in DKD was reproduced in an independent single-nucleus atlas.

These observations are relevant to how GLP-1 receptor agonists act in the diabetic kidney. Most DKD participants had kidney function within the range enrolled in FLOW, in which semaglutide slowed kidney disease progression [5], so the biopsies represent the stage at which treatment is usually started. At this stage, both smooth muscle cellularity and the fraction of cells carrying receptor transcripts were already reduced. Most of the deficit in receptor-carrying cells per arteriole, however, reflected lower receptor detection in the cells that remained rather than loss of the cells themselves, so the arteriolar target of these drugs was still present. A reduction in receptor expression within surviving cells may be more amenable to treatment than loss of the cells, which supports treatment at this stage. In mice, semaglutide lowers blood pressure through the smooth muscle receptor [12], and in REMODEL the intimal area of the most diseased arterioles was smaller after a year of treatment [13]; whether arteriolar responses depend on the receptor remaining at baseline can be tested in paired biopsies from treatment trials. Whole-kidney *Glp1r* downregulation has been reported in diabetic mice [14], and our measurements place that loss within the vascular wall of the human kidney. GLP-1 receptor activation also induces renin expression along the arteriolar tree in mice [11]. In our sections, renin-associated cells rarely carried *GLP1R* transcripts, and detection was concentrated in arteriolar smooth muscle, consistent with antibody localization [8].

The loss of smooth muscle cells connects the molecular data to the classical lesion. Arteriolar hyalinosis has long been described as the replacement of smooth muscle by hyaline material [1]. In keeping with this, walls with more PAS-dense acellular matrix contained fewer smooth muscle cells in every diagnosis, adjustment for recorded hyalinosis attenuated the diabetic deficit, and in an independent cohort smooth muscle content fell with each hyalinosis grade. The lost cells were mainly contractile cells without matrix gene expression, and higher matrix scores were confined to smooth muscle rather than wall fibroblasts or endothelium. Vessels with fewer than three smooth muscle cells were more frequent in DKD and lay outside the primary set, so the measured deficit excludes the most depleted vessels. Pericytes and perivascular fibroblasts [15] and Gli1-positive progenitors [16] contribute to matrix in experimental fibrosis, and their share cannot be separated in these sections.

The other selective changes point in different directions. *GRIA2* is expressed by human arterial smooth muscle and decreases after arterial injury in rats [17], and in cultured human smooth muscle cells it promotes proliferation and migration [18]. Lower *GRIA2* alongside fewer smooth muscle cells is consistent with a reduced capacity to maintain or replace the arteriolar smooth muscle layer, and *GRIA2* was also lower in KPMP. *PRDM6*, which promotes the proliferative gene program in smooth muscle cells [19], was lower than in other glomerular diseases in our cohort but higher in DKD in KPMP, so this change was not consistent between cohorts. By contrast, *P2RX1*, which mediates pressure-dependent afferent arteriolar constriction [20], was preserved in the remaining diabetic cells. Lower wall *REN* was not explained by RAS inhibitors and is in line with the hyporeninemic state of DKD [21].

The comparison with other glomerular diseases places these changes within diabetes rather than within kidney injury in general. IgAN, MCD and MN cover a wide range of proteinuria and kidney function, yet none showed significant reductions in arteriolar *GLP1R*, *GRIA2* or *PRDM6* relative to donors. The diabetic difference also survived adjustment for eGFR, proteinuria and hypertension, and participants with hypertension but without diabetes retained *GLP1R* detection in arteriolar smooth muscle. Given that hyperglycemia lowers receptor expression in beta cells [22] and that receptor loss accompanies diabetic injury in glomerular endothelium [23], these findings suggest that the diabetic milieu itself shapes the population of smooth muscle cells carrying receptor transcripts. This study has several limitations. Receptor protein was not measured, and detection in only 1% to 2% of donor wall cells makes the transcript estimates sparse. The cross-sectional design cannot order cell loss and transcriptional change. Arteriolar walls were outlined by automated segmentation without pathologist review, and arterioles were not distinguished from small arteries by morphology. Batches, eGFR and hypertension were imbalanced, and donor biopsies carried procurement signatures. Biopsies from people with diabetes but without kidney disease were not available. The panel lacked several smooth muscle genes, including *MYH11*, *CNN1*, *DES* and *TAGLN*.

In human DKD, the arteriolar wall loses contractile smooth muscle cells and the cells that remain carry less *GLP1R*, a pattern not shared by other glomerular diseases. These changes link the classical arteriolar lesion of diabetic nephropathy to the cellular target of GLP-1 receptor agonists at the disease stage in which these drugs are used.

## 4 Materials and Methods

### 4.1 Study design

This cross-sectional study asked whether arteriolar smooth muscle cells and their *GLP1R* expression are preserved in human DKD. All KORNERSTONE participants whose kidney biopsy sections had undergone single-cell-level spatial transcriptomics were included: participants with DKD, participants with IgAN, MCD, MN or FSGS as disease comparators, and living kidney donors. No sample size calculation was performed. One primary section per participant was analyzed, and the participant was the unit of analysis. Arterioles were identified by the same morphological and transcriptomic criteria in all sections, and these criteria did not use diagnosis. The principal outcomes were smooth muscle density and *GLP1R* positivity in arteriolar walls in DKD compared with other glomerular diseases; mural-gene expression was a secondary outcome, and the KPMP single-nucleus atlas served as an external cohort. Participants were grouped by clinical and pathological diagnosis, and randomization and blinding were not applicable.

### 4.2 Participants

Kidney biopsies were obtained from 154 participants in KORNERSTONE, a prospective Korean kidney biopsy cohort and biobank [24]: 28 with DKD, 60 with IgA nephropathy (IgAN), 25 with minimal change disease (MCD), 13 with membranous nephropathy (MN) and four with focal segmental glomerulosclerosis (FSGS), together with 24 living kidney donors who underwent zero-time biopsy at transplantation. One primary section per participant was analyzed. Among the 104 disease participants with linked pre-biopsy medication records, including 23 of 28 with DKD, none was receiving a GLP-1 receptor agonist. Estimated glomerular filtration rate (eGFR) was calculated with the 2021 CKD-EPI creatinine equation [25], and hypertension and diabetes were recorded at biopsy; for living donors, creatinine, proteinuria, hypertension and diabetes were taken from the pre-donation evaluation. The study was approved by the Institutional Review Board of Seoul National University Hospital (approval No. 2504-168-1637), and all participants gave written informed consent after the nature and possible consequences of the study had been explained.

### 4.3 Spatial transcriptomics and morpho-transcriptomic arteriole definition

Sections underwent single-cell-level spatial transcriptomics with the Xenium Prime 5K panel and a 100-gene add-on [26], and registered PAS histology was available for 146 participants.

Arterioles were identified by a morpho-transcriptomic definition that combined the morphology of each structure on PAS with the transcriptomic identity of the cells it contained (Fig. 1a; fig. S1). A deep-learning segmentation model outlined tissue structures on the PAS section (Supplementary Materials and Methods). The morphological criteria required an enclosed central lumen of low optical density, a lumen-to-outer diameter ratio of at least 0.2 and no overlap with a glomerulus; the wall was the area between the outer contour and the lumen. The transcriptomic criteria were applied to the registered cells: structures in which glomerular or tubular cells predominated, or with tubular cells in the lumen or more than two in the wall, were excluded, and at least three arteriolar smooth muscle cells had to lie within the wall. Structures meeting both sets of criteria were analyzed as arterioles and small arteries, and outer-diameter tertiles defined three calibers. The outline provides the wall area and places each cell inside or outside the wall, and the cells within the wall provide the counts and expression measured in that arteriole.

### 4.4 Wall measurements

Smooth muscle cells were counted per wall area and per 100 µm of outer perimeter, and all wall nuclei and *ACTA2* -positive cells were also counted irrespective of annotation. Each smooth muscle cell with at least one detected nuclear transcript was classed by whether it carried at least one nuclear transcript of the contractile-score genes and of the matrix genes *COL1A1*, *COL1A2*, *FN1* and *TNC*. The PAS-dense acellular fraction of each wall was measured by color deconvolution. Gene scores summarized contractile, mural-identity, matrix and proliferation expression.

### 4.5 *GLP1R* detection

Detection and pseudobulk expression analyses used mural-lineage smooth muscle cells within the arteriolar wall. Positivity required at least one nuclear *GLP1R* transcript. Negative-control probes, codewords and intergenic probes were processed identically and converted to false-positive rates expected for *GLP1R*. A cell mixed model estimated ratios of detection rates adjusted for nuclear transcripts per cell (Supplementary Materials and Methods). Proximal-tubule, endothelial and intergenic-probe positivity served as negative-control outcomes.

### 4.6 Gene expression

Counts from arteriolar smooth muscle cells were summed by participant, retaining the participant as the biological replicate [27]. Pseudobulk counts underwent trimmed mean of M-values normalization [28], mean–variance precision weighting [29] and empirical-Bayes moderation [30]. Mural genes were the 118 panel genes with higher mean expression in mural cells than in any other lineage, selected without diagnosis labels, and each non-diabetic disease was also compared with donors. The same participant pseudobulk was formed for fibroblasts and endothelial cells in the wall. Gene Ontology [31] gene-set enrichment [32] used rankings adjusted for transcript yield. Immediate-early and glucocorticoid-response scores quantified procurement effects in donor biopsies.

### 4.7 External cohort

Vascular smooth muscle in the Kidney Precision Medicine Project (KPMP) single-nucleus atlas [33] was analyzed by participant-level pseudobulk, comparing DKD with reference kidneys (table S6). Arteriolar hyalinosis grades were taken from the atlas pathology annotations.

### 4.8 Statistical analysis

Smooth muscle density and wall *GLP1R* positivity in DKD versus other glomerular diseases were the principal outcomes, with Holm adjustment [34]; mural-gene tests used Benjamini–Hochberg false discovery rate control within the 118 genes [35]. Wall models included diagnosis, caliber and a participant random intercept. Other glomerular diseases were weighted by participant frequency, and each diagnosis was also reported separately. Adjusted models added age, sex and batch, and further models added eGFR, proteinuria, hypertension or recorded hyalinosis. Comparisons with donors are reported descriptively. Tests were two-sided, and estimates are given with 95% CIs. The Supplementary Materials and Methods provide model equations and resampling definitions.

## Supporting information

Data file S1. Arteriole-level and gene-level summary data underlying the figures and tables.

## Acknowledgments

The biospecimens and data used for this study were provided by the Biobank of Seoul National University Hospital, a member of the Korea Biobank Network, and by the Korea National Institute of Health (KNIH) research project (project No. KBN5_A03).

## Author contributions

S.G.H. performed the spatial and statistical analyses and drafted the manuscript. M.K. developed the kidney structure segmentation model and produced the PAS outlines. D.K. performed cell-type annotation of the spatial transcriptomic data. J.H.J. and J.S.L. contributed to data generation and reviewed the final manuscript. H.K., A.K., J.-I.O., J.L., M.W.K., Y.S., H.L., S.S.H., Y.K., J.K., J.H.L., J.P.L., J.I.P., J.T.P., J.N.A., K.-C.M. and H.J.K. contributed to participant enrollment and data generation and reviewed the final manuscript. Y.-G.K. supervised image analysis and segmentation. S.P. conceived the study, curated clinical data, drafted the manuscript and supervised the analyses. D.K.K. established the cohort and supervised the study. All authors revised the manuscript and approved the final version.

## Data and materials availability

Individual-level spatial transcriptomic and clinical data are not publicly available and cannot be deposited in a public or controlled-access repository. The biopsy tissue was obtained with written broad consent for research use, but this consent and the approval of the Institutional Review Board of Seoul National University Hospital do not cover public release of individual-level data or their transfer outside Korea. De-identified data can be made available to researchers through a collaboration agreement with the KORNERSTONE investigators, subject to separate approval for each study by the Institutional Review Board and the Data Review Board of Seoul National University Hospital; requests should be addressed to the corresponding authors. Arteriole-level and gene-level summary data underlying the figures and tables are provided in data file S1. KPMP single-nucleus data are publicly available from the KPMP data portal (https://atlas.kpmp.org). Code for the kidney structure segmentation model is available from the corresponding authors upon request. The PAS images and manual annotations used to train the model, and the trained model weights derived from them, are not publicly available because the images are institutional biopsy images used under institutional review board approval. Analysis code is deposited at Zenodo (https://doi.org/10.5281/zenodo.24021047) (access on request).

## Supplementary Materials

### Materials and Methods

#### Participants and clinical data

Clinical variables were taken from the deidentified clinical database of the cohort and from the KORNERSTONE case report form. Records were linked to Xenium sections by pathology number in an access-controlled environment, and identifiers were not retained in analysis files.

Section-to-participant linkage was confirmed against restricted sample metadata for all 60 participants with IgAN. Matching by baseline laboratory eGFR, integer age and sex uniquely linked all 130 disease participants. Donor sex was taken from donor records, and donor age was calculated in completed years from the birth date to the date of donation. Donor creatinine, urine protein-to-creatinine ratio (UPCR), urine dipstick albumin, hypertension, diabetes and glycated hemoglobin were taken from the pre-donation evaluation in the electronic medical record, and donor eGFR was calculated from that creatinine with the 2021 CKD-EPI equation [S1] using donor age and sex. Creatinine was measured a median of 1 day before donation (range, 1 to 4 days) and UPCR a median of 98 days before donation (range, 22 to 462 days); urine dipstick albumin, measured 1 to 22 days before donation, was negative in 22 donors and trace in 2. No donor had diabetes (glycated hemoglobin, 4.7% to 5.8%).

In disease participants, hypertension and diabetes were recorded at biopsy. Pre-biopsy medication use was classified from case-report-form drug names in the 104 disease participants with available medication records.

#### Cell annotation

Cells were annotated following the cell-type hierarchy of the KPMP kidney atlas [S2]. Spatial transcriptomic data were integrated with KORNERSTONE single-nucleus RNA-sequencing data by scVI [S3] and clustered in the shared latent space. Clusters were assigned by canonical markers on the panel, and the single-nucleus profiles of the corresponding clusters were used for markers absent from the panel. Glomerular cells comprised podocytes (*NPHS1*, *NPHS2*, *PTPRQ*), parietal epithelial cells (*CFH*, *ALDH1A2*) and mesangial cells (*ITGA8*, *PIEZO2*, *POSTN*); tubular cells comprised proximal tubule (*LRP2*, *CUBN*, *SLC34A1*), injured proximal tubule (*VCAM1*, *HAVCR1*), loop of Henle (*SLC44A5*, *UMOD*, *SLC12A1*), distal nephron (*SLC12A3*, *CALB1*) and collecting duct cells (*AQP2*, *SLC4A1*, *SLC26A4*); and interstitial cells comprised endothelial cells (*PECAM1*, *EMCN*, *PLVAP*), mural cells (*ACTA2*, *MYH11*), fibroblasts (*DCN*, *C7*, *PDGFRA*), immune cells (*PTPRC*, *CD163*) and Schwann cells (*CDH19*, *SCN7A*). Renin-associated cells were identified by *REN* expression. Annotation did not use diagnosis, and the same labels were used for all analyses in this study.

#### Xenium processing and counts

Each section’s cell-feature matrix was matched to the analysis cell table by centroid. Nuclear counts comprised transcripts that were assigned to a cell, had a quality value of at least 20 and overlapped the segmented nucleus; all-assigned counts used the same rule without the nuclear criterion.

Technical control features were counted per cell with identical filters: 40 negative-control probes, negative-control codewords, unassigned codewords and intergenic genomic-control probes. Deprecated codewords were not used as background. Counts in each control category were converted to a *GLP1R*-equivalent rate, *λ* = 3 × count / (codewords × cells), because *GLP1R* is encoded by three codewords, and the expected number of false-positive cells among *n* cells was *n*(1 − exp(−*λ*)).

#### Automated segmentation of kidney structures

Kidney structures were outlined by an instance segmentation model built on Mask R-CNN [S4] with a ResNet-50 feature pyramid network backbone [S5, S6], initialized with weights trained on the COCO dataset [S7], to which convolutional block attention modules (CBAM) [S8] were added at each feature-pyramid level and before the box and mask heads (fig. S5a). The model took 512 × 512-pixel PAS tiles at 10× magnification (1.05 µm per pixel) and returned a mask for each structure; glomeruli, tubules and vessels were segmented as a single class, lumens were not delineated, and vessels were identified among the segmented structures after registration. For training, a trained annotator outlined glomeruli, tubules and vessels by hand in 94 patches of 512 × 512 pixels sampled at random from two PAS-stained whole-slide images of kidney biopsies outside the study cohort (3,268 structures), and the patches were split at random into training (60), validation (15) and test (19) sets. Both slides were obtained at Seoul National University Hospital, and their use was approved by its Institutional Review Board (approval No. H-2103-091-1205), which waived the requirement for informed consent. Training used flips, 90° rotations and color jitter, the Mask R-CNN losses with the box and mask terms weighted 3, the AdamW optimizer [S9] and cosine annealing with warm restarts [S10], and the weights with the highest validation mean average precision were kept. On the test patches, mask average precision at an intersection over union of 0.5 was 0.839, compared with 0.652 for Mask R-CNN without CBAM (fig. S5b). The model was applied to tiles of the whole PAS slides carrying the 146 sections with registered histology, with adjacent tiles overlapping by half and tiles with more than 20% tissue retained, and detections with a confidence score above 0.3 were pooled across the tiles of each slide. Duplicate detections from overlapping tiles were removed or merged according to their overlap relative to the area of the smaller detection, which approaches 1 when a partial detection lies within a complete one. For morphological cluster labels, each instance was taken as a 224 × 224-pixel image at 10× magnification, color-normalized by the Vahadane method [S11], masked to its contour and embedded with the UNI pathology foundation model [S12]; the embeddings were reduced to 32 dimensions with UMAP [S13] and grouped by k-means, both fitted on a random 10% of the instances from the 24 slides of the first batch. Sixteen clusters, chosen among 16, 32, 64 and 128 by the best agreement with the dominant cell type of each instance in the registered spatial data (adjusted mutual information), labeled every instance, and the glomerular cluster was the one whose instances contained, on average, at least 30% podocytes, mesangial cells and parietal epithelial cells.

Periodic acid–Schiff images were registered to cell coordinates by landmark alignment with tile-wise local correction, and contours were mapped to cell coordinates with the same registration. Within each contour, the lumen was the connected low-optical-density region containing the contour center, defined by an Otsu threshold on mean optical density within the contour at 1.05 µm per pixel. Contours were set aside if this lumen touched the outer contour along more than 5% of its boundary, if the lumen was smaller than 4 µm^2^ or if the wall had fewer than three pixels. Of 250,248 segmented contours intersecting the 146 sections, those centered outside the section, those in the glomerular cluster, those in which glomerular cells made up at least a quarter or tubular cells at least half of the enclosed cells (minimum three cells), those without smooth muscle or endothelial cells within 5 µm and those without an enclosed central lumen were set aside, leaving 9,722 vascular profiles.

#### Morpho-transcriptomic arteriole definition and wall measurements

The definition combined morphological criteria on the PAS contour (an enclosed central lumen, a lumen-to-outer equivalent-diameter ratio of at least 0.2, no glomerular overlap and exclusion of the glomerular morphological cluster) with transcriptomic criteria on the registered cells (no glomerular or tubular predominance among enclosed cells, smooth muscle or endothelial cells within 5 µm, no tubular cells in the lumen, no more than two tubular cells inside the outer contour and at least three smooth muscle cells in the wall).

Profiles with at least three smooth muscle (mural-lineage) cells whose centroids lay in the wall were analyzed as arterioles and small arteries, and outer-diameter thresholds of 23.42 and 35.74 µm, the tertiles of the candidate profiles, defined three calibers.

Wall cellularity was expressed per wall area and per 100 µm of outer perimeter and compared with linear mixed models that had diagnosis and caliber as fixed effects and a participant random intercept. The comparator was the mean of IgAN, MCD, MN and FSGS, weighted by the number of participants with each diagnosis. All nucleated wall cells were counted, including nuclei removed by transcript-count filtering in the raw segmentation; wall composition used low-resolution lineage annotations, and cells with at least one *ACTA2* transcript were counted irrespective of annotation.

Gene scores were mean gene-standardized log_2_ counts per million in wall-cell pseudobulk. The contractile score used *ACTA2*, *ACTG2*, *SMTN*, *MYLK*, *KCNMB1*, *SYNPO2* and *CSRP1* ; the mural-identity score *NOTCH3*, *PDGFRB*, *RGS5*, *MCAM* and *CSPG4* ; the synthetic matrix score *COL1A1*, *COL1A2*, *FN1* and *TNC* ; the proliferation score MKI67 and TOP2A; and the dedifferentiation and adhesion score KLF4, KLF5, VCAM1, CD44, MYH10 and THBS1. *MYH11*, *CNN1*, *DES*, *LMOD1* and *TAGLN* are not on the panel.

The PAS-dense acellular wall fraction was derived by optical-density color deconvolution with hematoxylin (0.644, 0.717, 0.267) and PAS (0.175, 0.972, 0.155) vectors and batch-specific Otsu thresholds. Thick-walled candidates with zero to two smooth muscle cells, which passed the same anatomical exclusions, had endothelial support and had a wall at least as thick as the donor median for the same caliber, were added only in sensitivity analyses.

#### Smooth muscle co-expression and wall matrix

Among smooth muscle cells with at least one nuclear transcript, contractile positivity required at least one transcript of *ACTA2*, *ACTG2*, *SMTN*, *MYLK*, *KCNMB1*, *SYNPO2* or *CSRP1*, and matrix positivity at least one transcript of *COL1A1*, *COL1A2*, *FN1* or *TNC*. Densities of the four combinations were counted per arteriole wall area and compared by linear mixed models with diagnosis and caliber as fixed effects and a participant random intercept. Matrix and contractile positivity were analyzed by cell-level complementary log–log mixed models with a log nuclear transcript offset and participant random intercept, with age, sex and batch added in adjusted models. Matrix scores were calculated from lineage-specific participant pseudobulk with at least ten cells of the lineage, using the same normalization and gene-standardization rule as the smooth muscle scores, and compared by participant regression with HC3 standard errors. Associations of smooth muscle density with PAS-dense acellular wall fraction, per ten percentage points, used linear mixed models with caliber and a participant random intercept, with diagnosis added in the pooled diagnosis-adjusted model and separate fits within diagnosis groups.

#### *GLP1R* models

Detection and pseudobulk expression analyses used mural-lineage smooth muscle cells assigned to the vascular wall. A wall smooth muscle cell was *GLP1R*-positive if it contained at least one nuclear *GLP1R* transcript. The primary model was a cell-level binomial generalized linear mixed model with a complementary log–log link, a participant random intercept and an offset for log nuclear transcripts per cell, fitted by maximum likelihood with adaptive Gauss–Hermite quadrature (15 nodes). Profile-likelihood intervals and a parametric bootstrap likelihood-ratio test (400 replicates) supplemented the Wald intervals, with lme4 providing reference likelihood fits.

Sensitivity analyses used participant beta-binomial models; a background-aware model in which each participant’s intergenic-probe *GLP1R*-equivalent rate entered as a fixed hazard; further adjustment for eGFR and hypertension; restriction to Batch 1 or to the largest caliber; comparison with participants with hypertension but without diabetes; and downsampling of comparator walls to the distribution of wall-cell numbers in DKD (2,000 replicates). *GLP1R*-positive cells per arteriole were decomposed into wall cells per arteriole and positive fraction on the log scale, with 5,000 participant bootstrap resamples. The same models were applied to *GLP1R* in proximal tubule, endothelium and all cells and to wall intergenic-probe positivity as negative-control outcomes.

For the disjoint localization display, renin-associated mural cells were assigned to a separate group before division of the remaining mural cells by their position inside or outside the PAS-defined arteriolar wall. This display contains 29,229 arteriolar, 225,415 other mural and 7,122 renin-associated cells. Primary detection and expression inference retains all 29,594 arteriolar smooth muscle cells, including 365 renin-associated wall cells, none positive for nuclear *GLP1R*. Analyses without PAS used cell-defined vessel profiles or all mural cells and participant pseudobulk expression, with 151 and 154 eligible participants, respectively.

#### Expression, receptors and gene sets

Participant pseudobulk counts were normalized by trimmed mean of M-values, weighted by the mean–variance trend and moderated by empirical Bayes [S14–S16]. Basic models contained diagnosis, and adjusted models added age, sex and batch. False discovery rates were controlled within the 118 panel genes with maximal mean expression in mural rather than other cell lineages, a filter computed from all cells without diagnosis labels. The vasoregulatory receptor set comprised 48 panel genes and was tested within its own family. Procurement scores were means of standardized immediate-early (*FOS*, *EGR2*, *EGR3*, *ATF3*, *NR4A1*) and glucocorticoid-response (*PER1*, *ZBTB16*) genes in wall cells, all cells and proximal tubule.

Gene-set enrichment used moderated statistics from models that included transcript yield, with GO biological-process and molecular-function sets restricted to the panel [S17, S18]; the two reported sets concern extracellular matrix structural constituents and the response to transforming growth factor beta.

#### External cohort

Vascular smooth muscle (subclass VSMC) in the KPMP single-nucleus atlas [S2] was obtained from the KPMP data portal and analyzed by participant-level pseudobulk, comparing DKD with reference kidneys. Participants needed at least ten smooth muscle nuclei to enter pseudobulk analyses, which left 15 participants with DKD and 22 reference participants. Pseudobulk counts were processed as in the discovery cohort. Positivity was compared by participant permutation (20,000) and cluster bootstrap (5,000), with exact intervals for pooled rate ratios. Mural-gene effects were compared with those of this study, DKD versus donors, for the 115 mural genes estimable in both datasets by Spearman correlation, as a descriptive measure without a *P* value.

For arteriolar hyalinosis, the smooth muscle fraction of each sample was log_2_{(*M* + 0.5)*/N* }, where *M* is the number of smooth muscle nuclei and *N* the number of all nuclei; the 0.5 correction retained samples without smooth muscle nuclei. A linear model regressed this fraction on the atlas arteriolar hyalinosis grade (0 to 3), with age, sex and DKD as covariates, in the 62 participants with a hyalinosis grade and known sex.

#### Statistical reporting

The participant was the unit of inference throughout. Two principal outcomes, smooth muscle density and wall *GLP1R* positivity in DKD versus other glomerular diseases, were Holm-adjusted; all other analyses were secondary or exploratory, with false discovery rates controlled within the families stated in the tables and legends [S19]. Wall models were also compared within slides that carried a DKD section. Confidence intervals are 95%, and tests are two-sided. Analyses used Python 3.12 (pandas, statsmodels, scipy, shapely, pyarrow) and R 4.5 (lme4, ggplot2, patchwork).

#### Wall geometry, densities and disease contrasts

For profile *v* in participant *j*, let 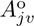 and 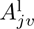 be the outer and luminal areas in *µ*m^2^, 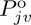 the outer perimeter in *µ*m and *M_jv_* the number of smooth muscle cells assigned to the wall. The equivalent diameters, wall area and two cellularity measures were

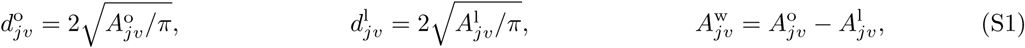

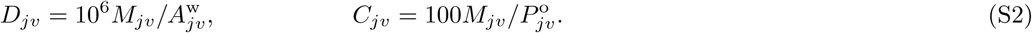

Thus *D_jv_* is cells/mm^2^ of wall and *C_jv_* is cells per 100 µm of outer perimeter. The lumen-to-outer diameter exclusion used 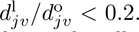 *<* 0.2. Primary profiles met all anatomical exclusions and contained at least three arteriolar smooth muscle cells. Endothelial-supported thick-wall profiles with zero to two smooth muscle cells entered sensitivity analyses, with the primary caliber thresholds held fixed.

For a continuous wall outcome, the basic model was

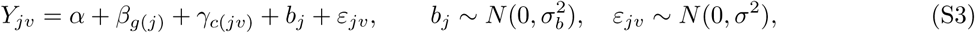

where *g*(*j*) denotes diagnosis and *c*(*jv*) caliber. Age, sex and batch were added in the adjusted model; models incorporating kidney function, proteinuria or pathology used the same outcome and comparison. If *n_g_* participants with wall measurements had other diagnosis *g*, its weight was *w_g_* = *n_g_/∑_h∈_*_O_ *n_h_*, for O = {IgAN, MCD, MN, FSGS}. The DKD contrast was

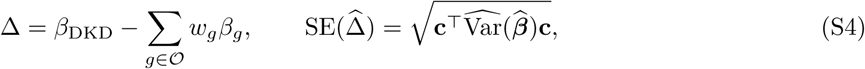

with the corresponding contrast vector **c**. On log scales, exponentiation gives a ratio; on the log_2_ expression scale, the estimate is a log_2_ fold change. This weighted disease contrast differs from pooling every comparator cell into a single sample. Participant bootstrap resampling retained diagnosis strata and each sampled participant’s vessels together. Slide-blocked comparisons used within-slide DKD and other-glomerular-disease measurements.

#### Participant-specific assay background

Let *C_jk_* be control transcript counts for category *k* in participant *j*, *K_k_* the number of codewords in that category and *N_j_* the number of sampled cells. Since the target uses three codewords, the section-level target-equivalent control intensity was

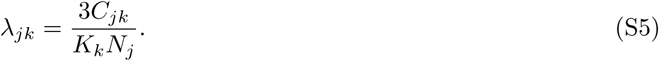

Technical backgrounds were scaled to wall cells by a category-specific pooled observed-to-expected wall ratio. Specifically, if 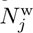 is the wall-cell count and 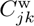 the wall-control count,

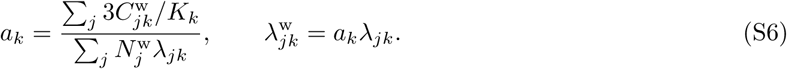

The sum of the four technical categories provided one background definition. Empirical proximal-tubule and endothelial backgrounds used each participant’s observed positive-cell fraction 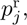, converted to intensity as 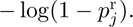 The wall-scaled versions multiplied this intensity by the pooled wall-to-reference intergenic-control rate ratio. The proximal-tubule background was the most conservative definition. Deprecated codewords were tabulated but excluded from background estimation. Group-level background estimates are reported in table S4.

For wall background intensity 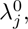, the expected false-positive probability and number were

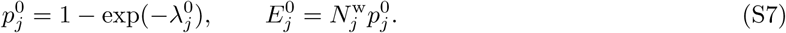

With *K_j_* observed positive wall cells, excess positivity was 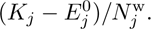 Negative excess estimates were retained to represent sampling uncertainty.

#### Cell-level detection likelihood and sensitivity estimands

For wall smooth muscle cell *i* of participant *j*, let *G_ij_* be its nuclear *GLP1R* count, *T_ij_* its total nuclear transcript count and *Z_ij_* = **1**(*G_ij_* ≥ 1). Cells with *T_ij_* = 0 were omitted from offset models. The primary conditional model was

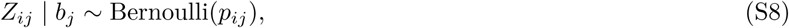

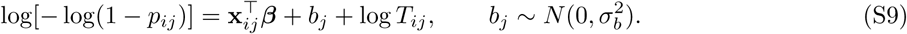

Here **x***_ij_* contains diagnosis and, in adjusted models, the specified participant covariates. Conditional on transcript yield and covariates, exp(**c**^T^***β***) is the ratio of positive-cell intensities per nuclear transcript. It is not the odds ratio from the logit sensitivity model. The participant likelihood integrated the within-participant Bernoulli contributions over the normal random intercept,

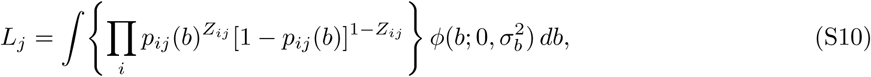

using adaptive Gauss–Hermite quadrature with 15 nodes. Profile-likelihood intervals and a 400-replicate parametric-bootstrap likelihood-ratio test supplemented Wald inference. Caliber, continuous diameter and an arteriole intercept nested within participant were alternative specifications. For background-aware detection, the target-associated intensity was added to the fixed participant background before calculating the positive probability:

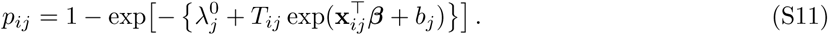

Participant beta-binomial models instead analyzed the number positive out of all wall cells and yielded odds ratios; Poisson and negative binomial count models yielded transcript-rate ratios with transcript-count offsets. Their denominators and effect scales were retained separately. Label permutations exchanged participants, not cells. Cell-count-matched downsampling held DKD counts fixed and repeatedly sampled comparator walls to the DKD wall-cell distribution.

#### Decomposition of positive-cell burden

Let *V_j_* be the number of included profiles, *N_j_* the sum of wall-cell counts over those profiles and *K_j_* their positive-cell sum. The burden identity was

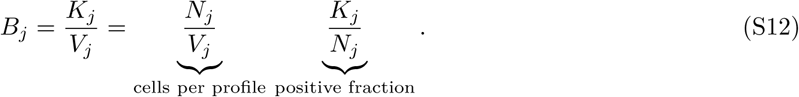

Profile-based sums were used consistently in both components. Within diagnosis, pooled component rates were calculated before taking the participant-frequency-weighted disease contrast. Consequently, on the log scale the basic contrast obeyed *δ_B_* = *δ_N_* + *δ_p_*. The contributions to the log burden difference were *δ_N_ /δ_B_* and *δ_p_/δ_B_*. Percentile intervals came from 5,000 diagnosis-stratified participant resamples, recomputing the rates, contrast and component shares each time. Shares can extend beyond zero or one when component effects oppose each other or the total is imprecise; they were not truncated.

#### Multiplicity

For the two principal outcomes, ordered *P* values *p*_(_*_r_*_)_ were Holm-adjusted as min{1, max*_ℓ_*_≤_*_r_*(*m* − *ℓ* + 1)*p*_(_*_ℓ_*_)_} with *m* = 2. Within each stated secondary or exploratory family, Benjamini–Hochberg values were *q*_(_*_r_*_)_ = min{1, min*_ℓ_*_≥_*_r_ mp*_(_*_ℓ_*_)_*/ℓ*}. These equations describe the reported correction families; the two principal outcomes use Holm correction. Full alternative-model estimates, support counts and correction scopes accompany the tables and data file S1.

### Supplementary Text

#### Smooth muscle density under alternative specifications

The deficit in DKD persisted within Batch 1 (−1,388 cells/mm^2^; −2,078 to −698) and after adjustment for eGFR and proteinuria (−1,338; −2,061 to −616), hypertension (−1,719; −2,467 to −970), global glomeru-losclerosis (−1,890; −2,577 to −1,203) or the PAS matrix fraction of the wall (−1,918; −2,547 to −1,290) (table S1). Adjustment for recorded arteriolar hyalinosis in 86 participants attenuated it (−1,416; −2,531 to −302), and its interval included zero once age, sex and batch were added (−855; −1,974 to 265). On the 28 slides that carried a DKD section, DKD walls were lower than same-slide comparators by 1,476 cells/mm^2^ (−2,327 to −624). Adding thick-walled profiles with zero to two smooth muscle cells enlarged the deficit (−2,440; −3,142 to −1,737). Against donors, smooth muscle was lower by 1,837 cells/mm^2^ (−2,619 to −1,054) and by 1.48 cells per 100 µm of perimeter (−2.30 to −0.66).

#### *GLP1R* detection under alternative specifications

Relative to other glomerular diseases, the ratio of *GLP1R* detection rates was 0.33 (0.17 to 0.66) in the basic model, 0.36 (0.18 to 0.69) after adjustment for age, sex and batch, and 0.36 (0.15 to 0.85) after further adjustment for eGFR and hypertension in 101 participants (table S4). Participant beta-binomial models gave odds ratios of 0.41 (0.21 to 0.78) and 0.40 (0.20 to 0.77), and the background-aware model with intergenic probes gave 0.32 (0.15 to 0.70). The reduction was present within Batch 1 (0.28; 0.13 to 0.60), in the largest caliber (0.35; 0.18 to 0.71) and against 23 participants with hypertension but without diabetes (0.34; 0.13 to 0.91). Negative-control outcomes did not differ: proximal-tubule positivity (odds ratio, 0.94; 0.70 to 1.26), endothelial positivity (0.87; 0.63 to 1.21) and wall intergenic-probe positivity (0.69; 0.46 to 1.06). Without PAS outlines, *GLP1R* expression was lower in Xenium-defined vessel profiles (ratio, 0.62; 0.46 to 0.85) and in all mural cells (0.55; 0.34 to 0.88).

---

**Fig. S1.**
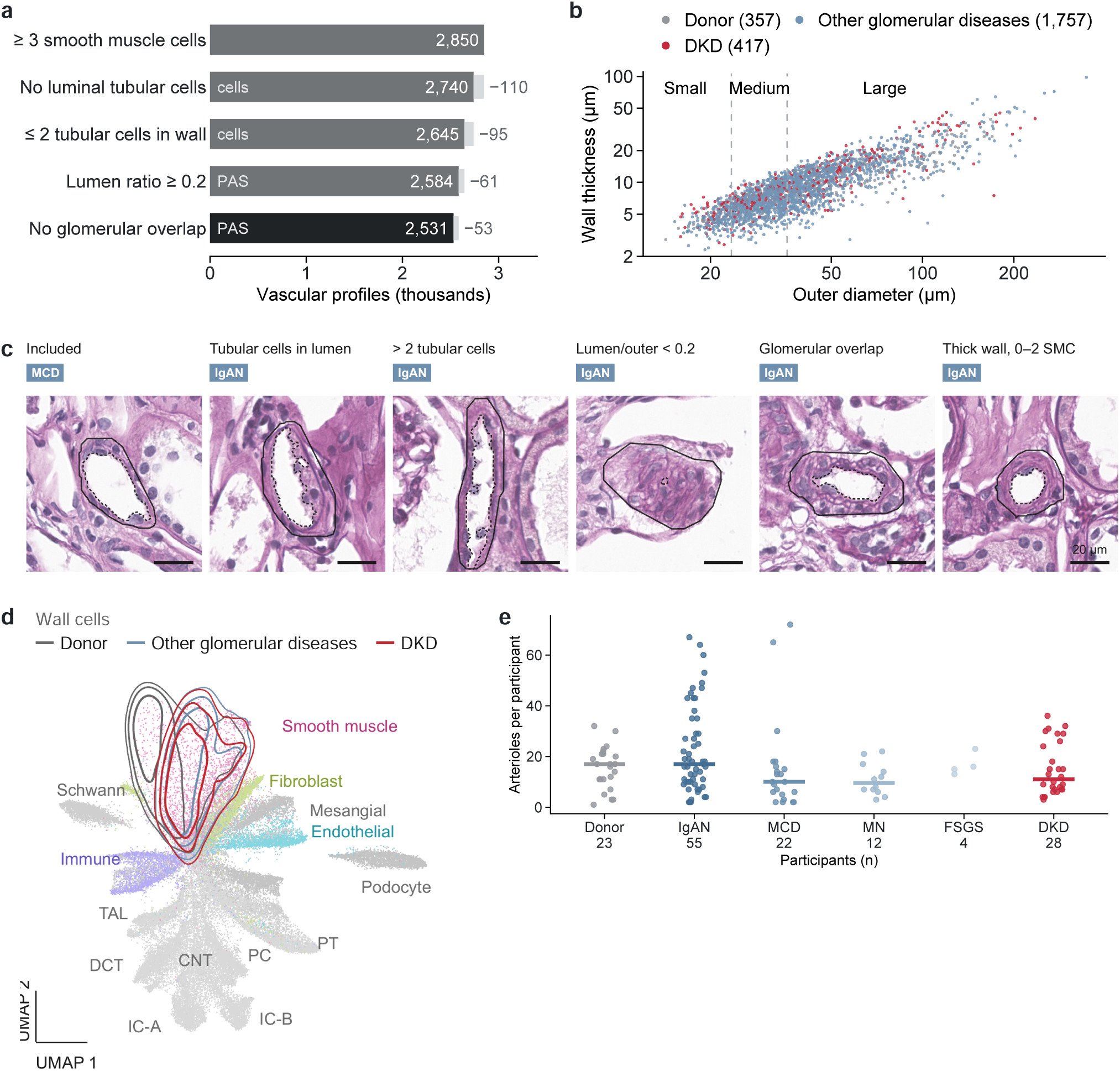
Morpho-transcriptomic arteriole definition. (a) Sequential exclusions reduced 2,850 profiles with at least three smooth muscle cells to 2,531 arterioles and small arteries in 144 participants; each step is marked as a morphological (PAS) or transcriptomic (cell) criterion, and counts are printed beside the bars. (b) Outer equivalent diameter versus wall thickness; dashed boundaries at 23.42 and 35.74 µm define small, medium and large calibers. (c) Six PAS examples showing an included profile and the indicated exclusion or thick-wall categories. Solid outlines mark outer contours and dashed outlines the lumen; scale bars, 20 µm. (d) Embedding of all cells with marker-defined identities; contours show arteriolar wall cells in donors, other glomerular diseases and DKD. (e) Eligible arterioles per participant. Points are participants and bars medians; FSGS is shown as points alone. Primary profile totals were 357 donor, 1,220 IgAN, 338 MCD, 132 MN, 67 FSGS and 417 DKD. The full cohort contained 5,195,294 cells from 154 participants; PAS was registered for 146.

**Fig. S2.**
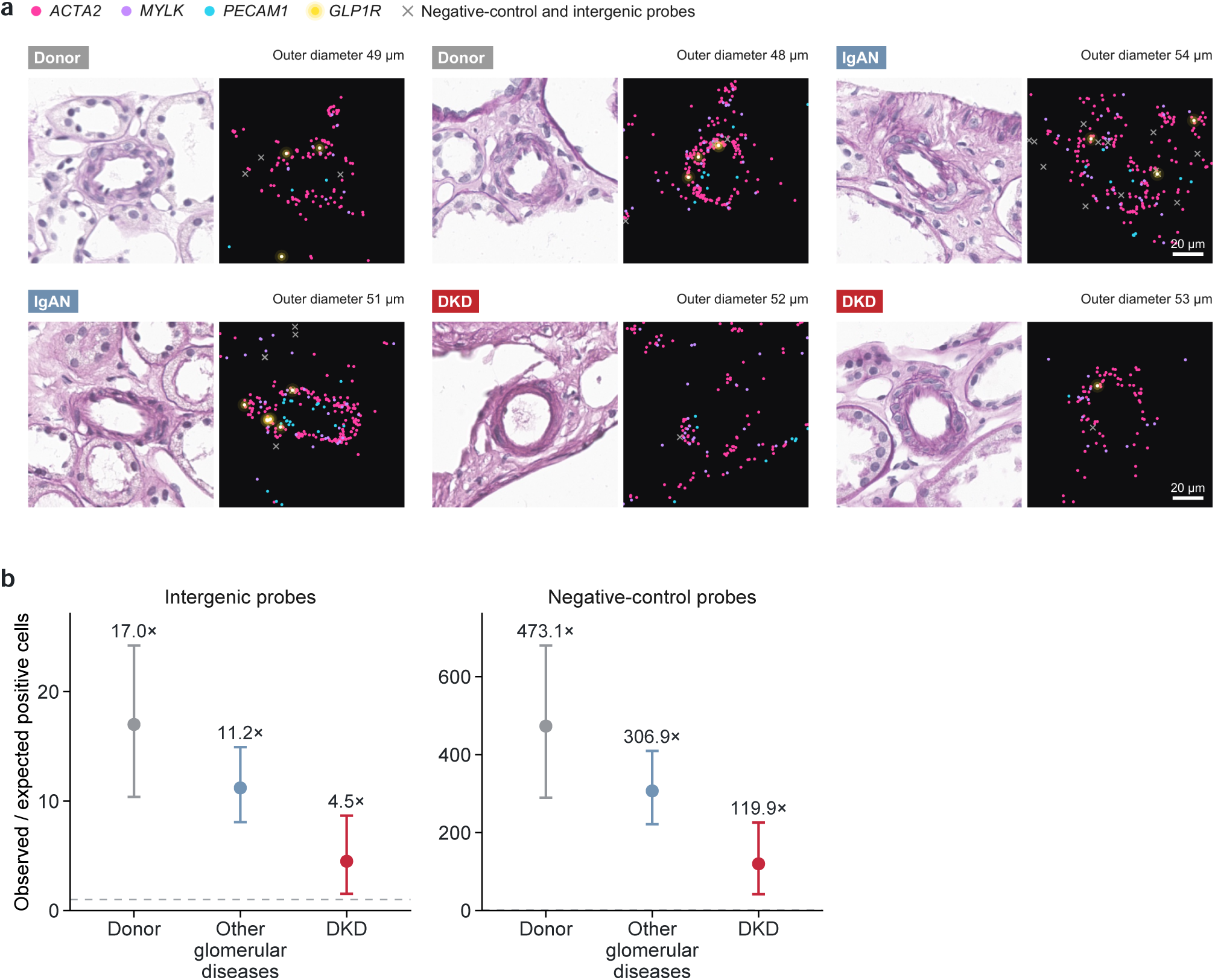
*GLP1R* detection. (a) Six representative arteriolar fields with PAS and nuclear transcript calls in the same physical field. *ACTA2*, *MYLK*, *PECAM1* and *GLP1R* calls are colored as indicated, and crosses mark negative-control and intergenic probe calls. Field identifiers and selection rules are given in the Representative fields worksheet of data file S1. Scale bars, 20 µm. (b) Observed-to-expected positive arteriolar smooth muscle cells with intergenic-probe and negative-control-probe backgrounds. Points are pooled group ratios with 95% CIs from 5,000 diagnosis-stratified participant resamples; dashed lines mark a ratio of one. Counts, background definitions and model estimates are in tables S3 and S4.

**Fig. S3.**
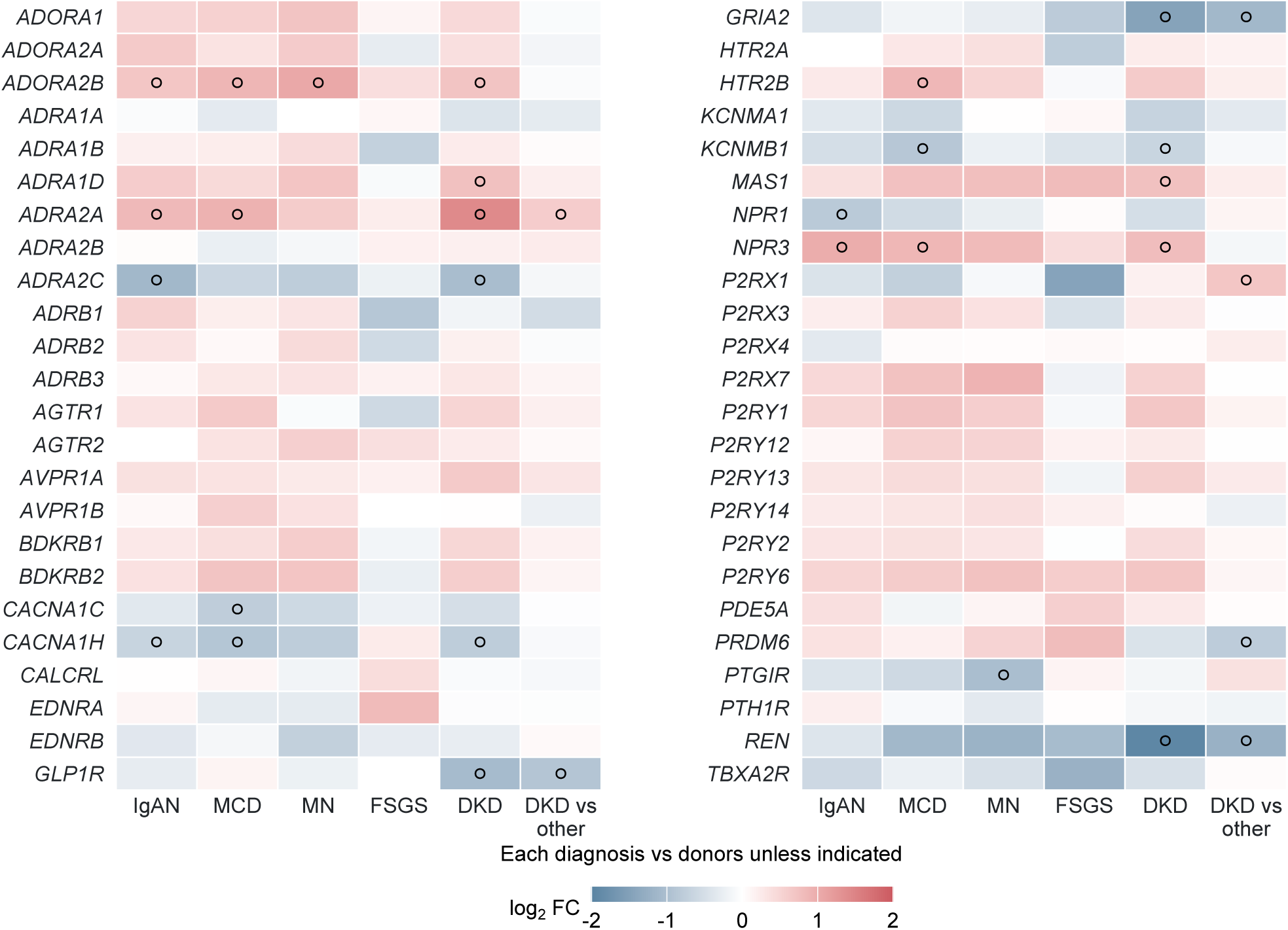
Vasoregulatory receptor panel. All 48 vasoregulatory panel genes, arranged alphabetically in two blocks, from participant pseudobulk. Columns compare each disease with donors and, in the final column, DKD with the participant-weighted mean of other glomerular diseases. Tiles show log_2_ fold change; open circles mark *q* < 0.05 within the 48-gene receptor family.

**Fig. S4.**
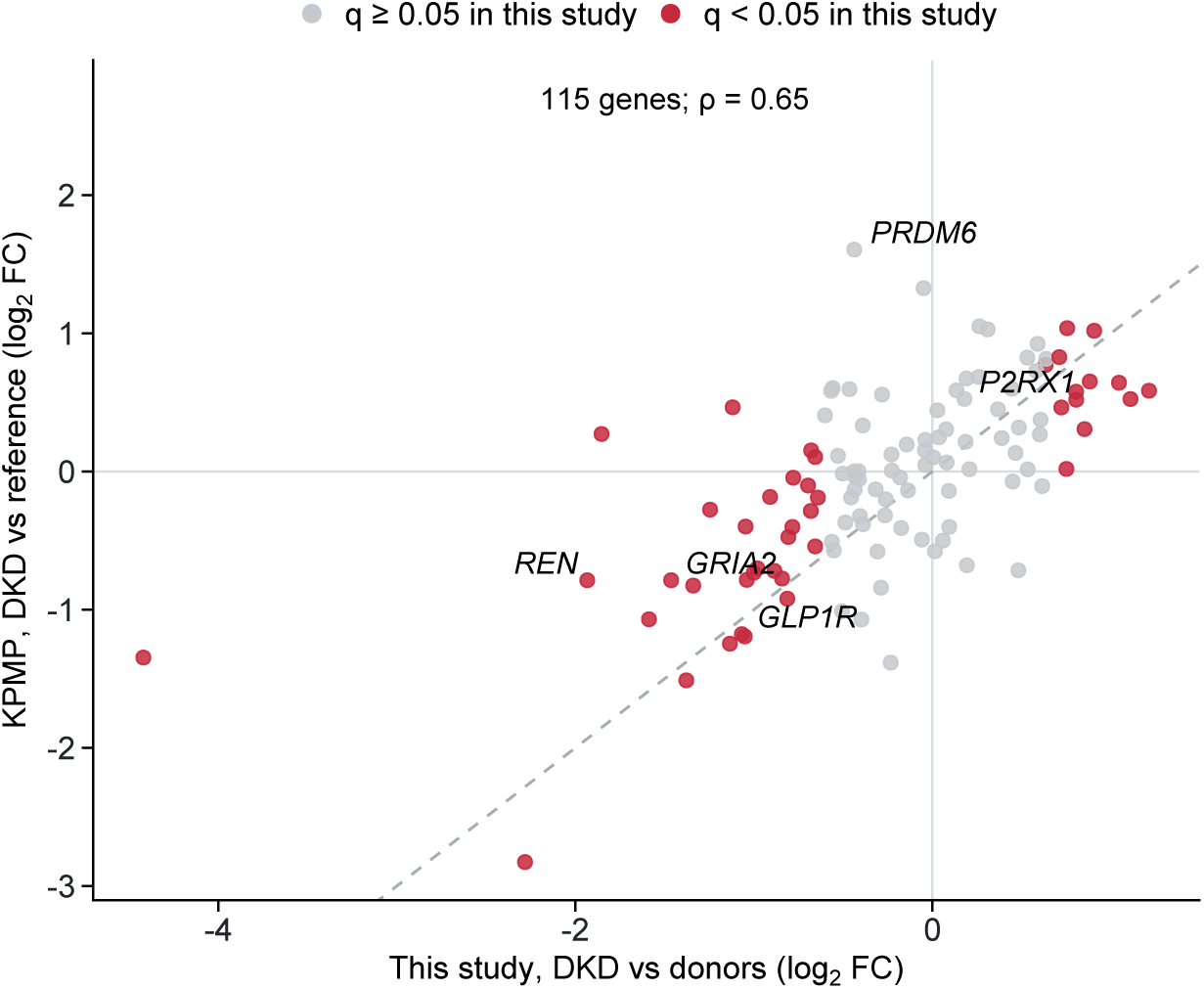
Mural-gene effects in this study and KPMP. Log_2_ fold changes for the 115 mural genes estimable in both datasets: this study, DKD versus donors (horizontal axis), and KPMP vascular smooth muscle, DKD versus reference kidneys (vertical axis). Each point is a gene; red points have *q* < 0.05 within the 118 mural genes in this study, and the five altered genes are labeled. The dashed line marks equal effects. The Spearman correlation is 0.65. The KPMP source population is described in table S6.

**Fig. S5.**
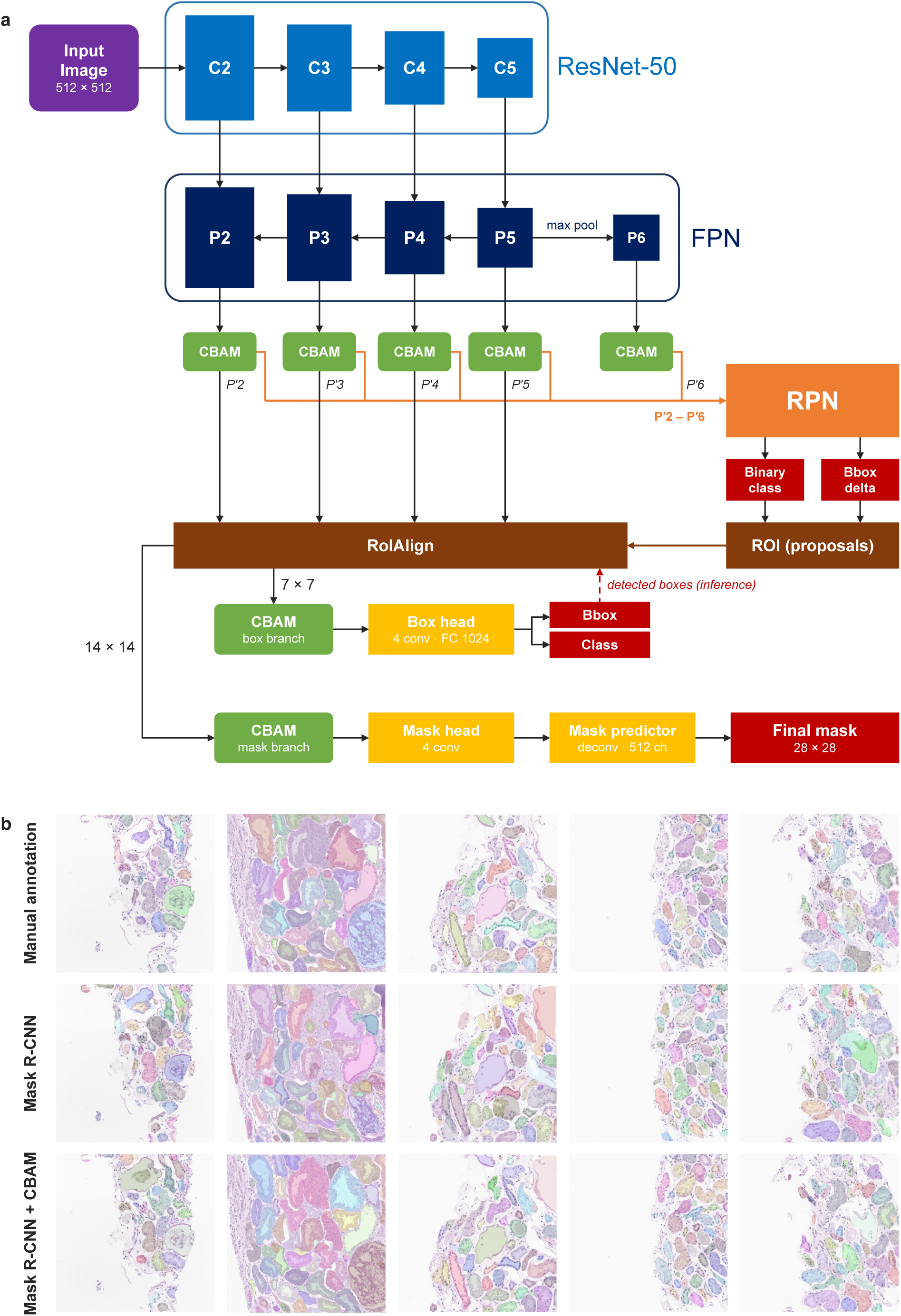
Kidney structure segmentation model. (a) Architecture. A ResNet-50 feature pyramid network computes feature maps P2–P6 from each 512 × 512-pixel PAS tile, and a convolutional block attention module (CBAM) refines each map. The refined maps Pr2–Pr6 enter the region proposal network, and region-of-interest alignment pools features for each proposed region from Pr2–Pr5, at 7 × 7 for the box branch and 14 × 14 for the mask branch. A further CBAM in each branch refines the region features before the box head, which returns the class score and box of each structure, and the mask head, which returns its mask. At inference, the mask branch uses the detected boxes (dashed arrow). (b) Manual annotation (top row) and predictions of Mask R-CNN (middle row) and of Mask R-CNN with CBAM (bottom row) on five test patches of 512 × 512 pixels at 10× magnification, one patch per column; colors distinguish individual structures.

**Supplementary Table S1.** Smooth muscle cellularity under alternative specifications.

| Analysis | *n* (DKD) | Basic difference (95% CI) | *q* | Adjusted difference (95% CI) | *q* |
|---|---|---|---|---|---|
| Batch 1 only | 95 (24) | −1,388 (−2,078 to −698) | 1.36 × 10<sup>−4</sup> | −1,191 (−1,848 to −534) | 7.56 × 10<sup>−4</sup> |
| + eGFR and log UPCR | 120 (27) | −1,338 (−2,061 to −616) | 4.15 × 10<sup>−4</sup> | −1,051 (−1,772 to −330) | 0.006 |
| + hypertension | 101 (24) | −1,719 (−2,467 to −970) | 1.35 × 10<sup>−5</sup> | −1,316 (−2,088 to −545) | 0.001 |
| + global glomerulosclerosis | 86 (23) | −1,890 (−2,577 to −1,203) | 2.40 × 10<sup>−7</sup> | −1,418 (−2,117 to −719) | 2.03 × 10<sup>−4</sup> |
| + recorded hyalinosis | 86 (20) | −1,416 (−2,531 to −302) | 0.014 | −855 (−1,974 to 265) | 0.152 |
| + PAS matrix fraction | 144 (28) | −1,918 (−2,547 to −1,290) | 9.67 × 10<sup>−9</sup> | −1,217 (−1,836 to −599) | 2.94 × 10<sup>−4</sup> |
| Same-slide comparators | 28 slides | −1,476 (−2,327 to −624) | — | — | — |
| Thick-walled profiles with 0–2 smooth muscle cells added | 144 (28) | −2,440 (−3,142 to −1,737) | 1.47 × 10<sup>−10</sup> | −1,853 (−2,567 to −1,139) | 5.29 × 10<sup>−6</sup> |
| Smooth muscle cells per mm<sup>2</sup> of wall, vs donors | 144 (28) | −1,837 (−2,619 to −1,054) | 9.77 × 10<sup>−6</sup> | −1,969 (−2,739 to −1,199) | 1.89 × 10<sup>−6</sup> |
| Smooth muscle cells per 100 µm of perimeter, vs donors | 144 (28) | −1.48 (−2.30 to −0.66) | 4.51 × 10<sup>−4</sup> | −1.48 (−2.31 to −0.66) | 7.64 × 10<sup>−4</sup> |
| PAS-dense matrix fraction, per 10 percentage points | Arterioles | Participants | Difference (95% CI),<br>cells/mm<sup>2</sup> | *P* |  |
| All arterioles | 2,531 | 144 | −793 (−934 to −653) | 2.27 × 10<sup>−28</sup> |  |
| DKD arterioles only | 417 | 28 | −579 (−860 to −297) | 5.58 × 10<sup>−5</sup> |  |

Difference in smooth muscle cells per mm<sup>2</sup> of wall, DKD minus other glomerular diseases, from linear mixed models with diagnosis, caliber and a participant random intercept, unless stated otherwise. *n* is participants, with participants with DKD in parentheses. Adjusted models add age, sex and batch. Hypertension and recorded pathology rows use participants with available records; hyalinosis was recorded by the pathologist on the case report form. The PAS matrix fraction is the PAS-dense acellular fraction of the wall. The lower block reports the matrix-fraction slope from caliber-adjusted linear mixed models with a participant random intercept, without diagnosis in the pooled model; its *P* values are unadjusted. The same-slide row compares DKD walls with other-glomerular-disease walls on the 28 slides that each carried one DKD section (design-based; *P* = 0.001; lower on 22 of 28 slides) and has no adjusted model or multiplicity-adjusted *q* value. Added thick-walled profiles passed the same anatomical exclusions, had endothelial support and a wall at least as thick as the donor median for the same caliber. Donor rows use the indicated units. *q* values are Benjamini–Hochberg values across the complete cellularity sensitivity family within each model, which is reported in full in data file S1. The donor rows use the seven tests of Table 2.

**Supplementary Table S2.** Gene-set enrichment, immediate-early responses and wall-lineage matrix scores.

| GO identifier | Gene set | Genes | NES, no yield adjustment | *q* | NES, yield-adjusted | *q* |  |
|---|---|---|---|---|---|---|---|
| GO:0005201 | extracellular matrix structural constituent | 70 | 2.25 | 0.001 | 2.44 | 0.032 |  |
| GO:0071559 | response to transforming growth factor beta | 85 | 1.51 | 0.107 | 2.00 | 0.034 |  |
| Compartment | Signature | Comparison | *n* | Basic difference (95% CI) | *q* | Adjusted difference (95% CI) | *q* |
| Arteriolar smooth muscle | IEG | DKD vs donors | 144 | −1.95 (−2.16 to −1.74) | 1.27 × 10<sup>−38</sup> | −1.90 (−2.12 to −1.68) | 1.14 × 10<sup>−34</sup> |
| Arteriolar smooth muscle | IEG | DKD vs other glomerular diseases | 144 | 0.07 (−0.07 to 0.21) | 0.405 | −0.02 (−0.16 to 0.12) | 0.742 |
| Arteriolar smooth muscle | GC | DKD vs donors | 144 | −1.85 (−2.17 to −1.52) | 1.28 × 10<sup>−20</sup> | −1.88 (−2.23 to −1.53) | 4.11 × 10<sup>−19</sup> |
| Arteriolar smooth muscle | GC | DKD vs other glomerular diseases | 144 | 0.05 (−0.27 to 0.36) | 0.773 | 0.09 (−0.24 to 0.41) | 0.742 |
| Matrix score by wall lineage | Participants (DKD) | Difference (95% CI) | *P* |  |  |  |  |
| Smooth muscle | 143 (28) | 0.64 (0.36 to 0.91) | 1.41 × 10<sup>−5</sup> |  |  |  |  |
| Fibroblasts | 72 (13) | 0.17 (−0.45 to 0.79) | 0.592 |  |  |  |  |
| Endothelium | 136 (27) | −0.06 (−0.27 to 0.16) | 0.614 |  |  |  |  |

DKD versus other glomerular diseases, basic participant-pseudobulk model. NES, normalized enrichment score; *q* values use the complete measured GO biological-process and molecular-function family. The adjusted ranking includes transcript yield. The gene universe contains 5,094 measured genes. Both sets pass the complete GO-family FDR threshold after yield adjustment. Score differences (95% CI) in mean gene-standardized log<sub>2</sub> CPM. Immediate-early genes (IEG): *FOS*, *EGR2*, *EGR3*, *ATF3*, *NR4A1*. Glucocorticoid-response genes (GC): *PER1*, *ZBTB16*. Each compartment is standardized across its participants. Basic participant-level HC3 models contain diagnosis; adjusted models add age, sex and batch. Other glomerular diseases are weighted by participant frequency. Benjamini–Hochberg correction is over both signatures and the two DKD contrasts within compartment, core-set definition and model. The complete spatial cohort has 154 participants and the eligible wall population 144. Donor procurement conditions differ from indication biopsy conditions; donor contrasts are descriptive. Matrix scores use the four matrix genes and at least ten cells per lineage per participant. The smooth muscle estimate here (0.64; 143 participants) uses this threshold, whereas the main-text estimate (0.61; 144 participants) includes every participant with eligible arteriolar smooth muscle. The lineage-score *P* values are unadjusted.

**Supplementary Table S3.** Participant wall smooth muscle cell and GLP1R count distributions.

| Diagnosis | *n* | Wall SMCs | Nuclear<br>transcripts | Positive<br>wall cells |
|---|---|---|---|---|
| Living donors | 23 | 218 (132–303) | 4 (0–6) | 4 (0–6) |
|  |  | Total 5,078 | Total 99 | Total 89 |
| Diabetic kidney disease | 28 | 126 (70–203) | 0 (0–1) | 0 (0–1) |
|  |  | Total 4,145 | Total 20 | Total 20 |
| IgA nephropathy | 55 | 221 (130–338) | 2 (0–2) | 2 (0–2) |
|  |  | Total 13,972 | Total 177 | Total 161 |
| Minimal change disease | 22 | 100 (34–316) | 2 (0–4) | 2 (0–4) |
|  |  | Total 3,909 | Total 68 | Total 60 |
| Membranous nephropathy | 12 | 106 (77–178) | 1 (0–1) | 1 (0–1) |
|  |  | Total 1,444 | Total 17 | Total 17 |
| Focal segmental glomerulosclerosis | 4 | 218 (152–328) | 2 (2–3) | 2 (2–3) |
|  |  | Total 1,046 | Total 9 | Total 9 |

Median (interquartile range) per participant and absolute total. Nuclear transcripts with quality score at least 20; eligible arteriolar smooth muscle cells; SMC, smooth muscle cell. Matched reference detection was 3,386/1,214,906 proximal-tubule cells and 1,104/589,600 endothelial cells. Wall detection was 356/29,594 cells.

**Supplementary Table S4.**
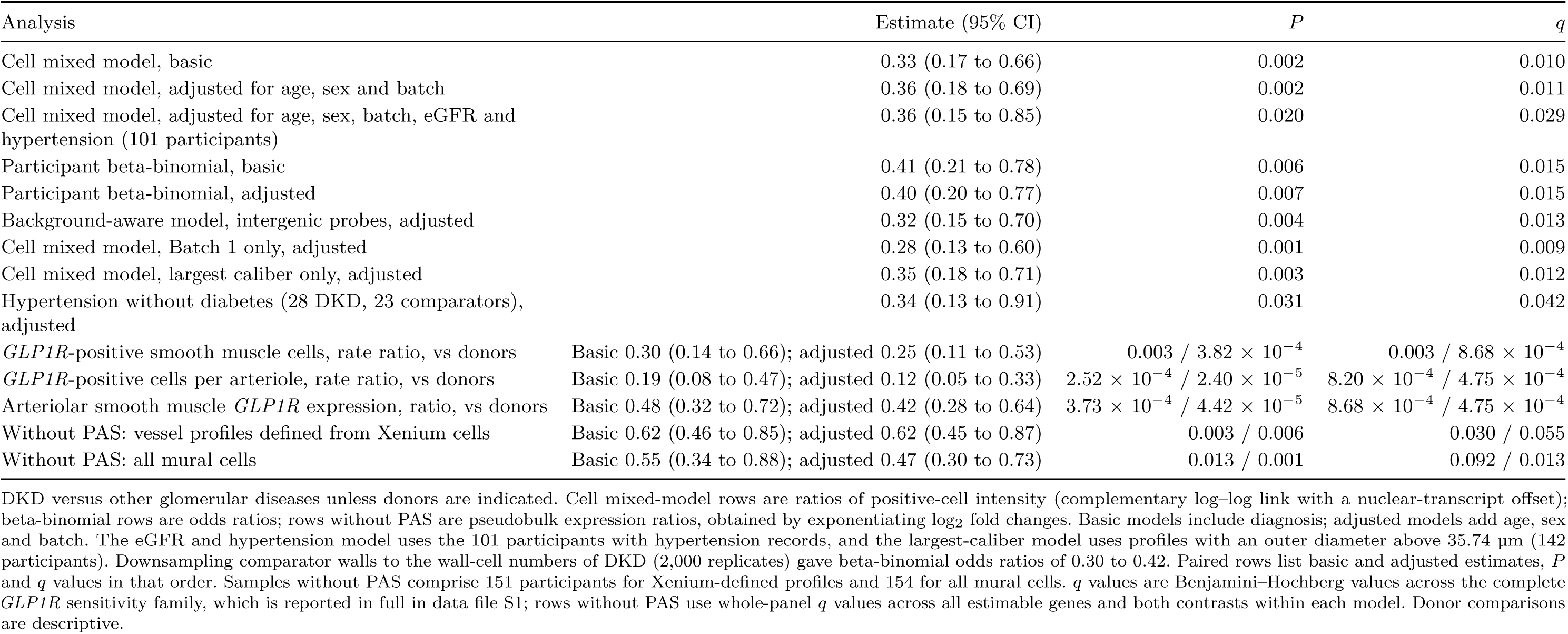
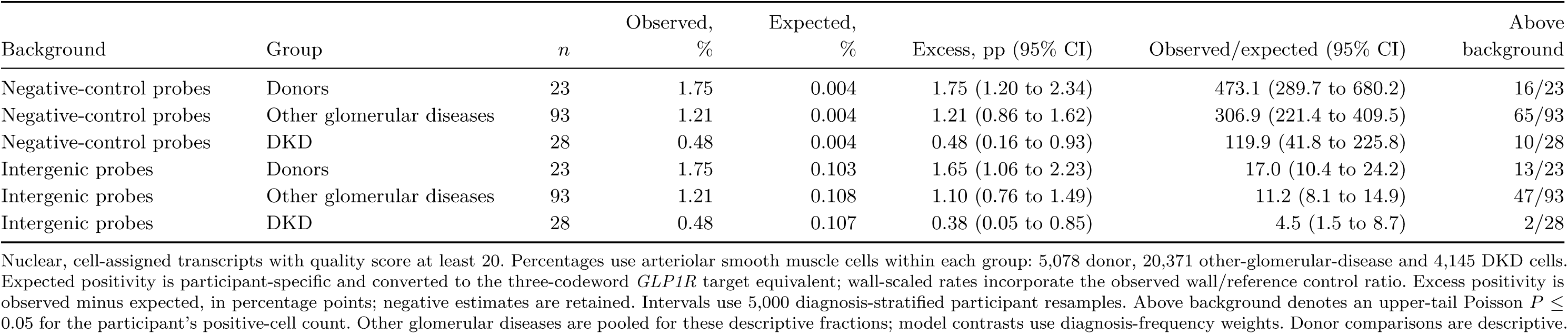
GLP1R sensitivity and assay background.

| Analysis | Estimate (95% CI) | *P* | *q* |  |  |  |  |
|---|---|---|---|---|---|---|---|
| Cell mixed model, basic | 0.33 (0.17 to 0.66) | 0.002 | 0.010 |  |  |  |  |
| Cell mixed model, adjusted for age, sex and batch | 0.36 (0.18 to 0.69) | 0.002 | 0.011 |  |  |  |  |
| Cell mixed model, adjusted for age, sex, batch, eGFR and hypertension (101 participants) | 0.36 (0.15 to 0.85) | 0.020 | 0.029 |  |  |  |  |
| Participant beta-binomial, basic | 0.41 (0.21 to 0.78) | 0.006 | 0.015 |  |  |  |  |
| Participant beta-binomial, adjusted | 0.40 (0.20 to 0.77) | 0.007 | 0.015 |  |  |  |  |
| Background-aware model, intergenic probes, adjusted | 0.32 (0.15 to 0.70) | 0.004 | 0.013 |  |  |  |  |
| Cell mixed model, Batch 1 only, adjusted | 0.28 (0.13 to 0.60) | 0.001 | 0.009 |  |  |  |  |
| Cell mixed model, largest caliber only, adjusted | 0.35 (0.18 to 0.71) | 0.003 | 0.012 |  |  |  |  |
| Hypertension without diabetes (28 DKD, 23 comparators), adjusted | 0.34 (0.13 to 0.91) | 0.031 | 0.042 |  |  |  |  |
| *GLP1R*-positive smooth muscle cells, rate ratio, vs donors | Basic 0.30 (0.14 to 0.66); adjusted 0.25 (0.11 to 0.53) | 0.003 / 3.82 × 10<sup>−4</sup> | 0.003 / 8.68 × 10<sup>−4</sup> |  |  |  |  |
| *GLP1R*-positive cells per arteriole, rate ratio, vs donors | Basic 0.19 (0.08 to 0.47); adjusted 0.12 (0.05 to 0.33) | 2.52 × 10<sup>−4</sup> / 2.40 × 10<sup>−5</sup> | 8.20 × 10<sup>−4</sup> / 4.75 × 10<sup>−4</sup> |  |  |  |  |
| Arteriolar smooth muscle *GLP1R* expression, ratio, vs donors | Basic 0.48 (0.32 to 0.72); adjusted 0.42 (0.28 to 0.64) | 3.73 × 10<sup>−4</sup> / 4.42 × 10<sup>−5</sup> | 8.68 × 10<sup>−4</sup> / 4.75 × 10<sup>−4</sup> |  |  |  |  |
| Without PAS: vessel profiles defined from Xenium cells | Basic 0.62 (0.46 to 0.85); adjusted 0.62 (0.45 to 0.87) | 0.003 / 0.006 | 0.030 / 0.055 |  |  |  |  |
| Without PAS: all mural cells | Basic 0.55 (0.34 to 0.88); adjusted 0.47 (0.30 to 0.73) | 0.013 / 0.001 | 0.092 / 0.013 |  |  |  |  |
| Background | Group | *n* | Observed, % | Expected, % | Excess, pp (95% CI) | Observed/expected (95% CI) | Above background |
| Negative-control probes | Donors | 23 | 1.75 | 0.004 | 1.75 (1.20 to 2.34) | 473.1 (289.7 to 680.2) | 16/23 |
| Negative-control probes | Other glomerular diseases | 93 | 1.21 | 0.004 | 1.21 (0.86 to 1.62) | 306.9 (221.4 to 409.5) | 65/93 |
| Negative-control probes | DKD | 28 | 0.48 | 0.004 | 0.48 (0.16 to 0.93) | 119.9 (41.8 to 225.8) | 10/28 |
| Intergenic probes | Donors | 23 | 1.75 | 0.103 | 1.65 (1.06 to 2.23) | 17.0 (10.4 to 24.2) | 13/23 |
| Intergenic probes | Other glomerular diseases | 93 | 1.21 | 0.108 | 1.10 (0.76 to 1.49) | 11.2 (8.1 to 14.9) | 47/93 |
| Intergenic probes | DKD | 28 | 0.48 | 0.107 | 0.38 (0.05 to 0.85) | 4.5 (1.5 to 8.7) | 2/28 |

DKD versus other glomerular diseases unless donors are indicated. Cell mixed-model rows are ratios of positive-cell intensity (complementary log–log link with a nuclear-transcript offset); beta-binomial rows are odds ratios; rows without PAS are pseudobulk expression ratios, obtained by exponentiating log<sub>2</sub> fold changes. Basic models include diagnosis; adjusted models add age, sex and batch. The eGFR and hypertension model uses the 101 participants with hypertension records, and the largest-caliber model uses profiles with an outer diameter above 35.74 µm (142 participants). Downsampling comparator walls to the wall-cell numbers of DKD (2,000 replicates) gave beta-binomial odds ratios of 0.30 to 0.42. Paired rows list basic and adjusted estimates, *P* and *q* values in that order. Samples without PAS comprise 151 participants for Xenium-defined profiles and 154 for all mural cells. *q* values are Benjamini–Hochberg values across the complete *GLP1R* sensitivity family, which is reported in full in data file S1; rows without PAS use whole-panel *q* values across all estimable genes and both contrasts within each model. Donor comparisons are descriptive.
Nuclear, cell-assigned transcripts with quality score at least 20. Percentages use arteriolar smooth muscle cells within each group: 5,078 donor, 20,371 other-glomerular-disease and 4,145 DKD cells. Expected positivity is participant-specific and converted to the three-codeword *GLP1R* target equivalent; wall-scaled rates incorporate the observed wall/reference control ratio. Excess positivity is observed minus expected, in percentage points; negative estimates are retained. Intervals use 5,000 diagnosis-stratified participant resamples. Above background denotes an upper-tail Poisson *P* ≤ 0.05 for the participant's positive-cell count. Other glomerular diseases are pooled for these descriptive fractions; model contrasts use diagnosis-frequency weights. Donor comparisons are descriptive.

**Supplementary Table S5.** Clinical covariate sensitivity of the five altered mural genes.

| Analysis | *n* | *GLP1R* | *GRIA2* | *PRDM6* | *REN* | *P2RX1* |
|---|---|---|---|---|---|---|
| Age, sex and batch (disease participants) | 121 | −0.90 (−1.37 to −0.43) | −1.08 (−1.53 to −0.63) | −0.65 (−1.11 to −0.20) | −1.13 (−1.84 to −0.42) | 0.57 (0.12 to 1.01) |
|  |  | *q* = 0.087 | *q* = 0.006 | *q* = 0.453 | *q* = 0.318 | *q* = 0.584 |
| + eGFR | 121 | −0.89 (−1.41 to −0.36) | −1.01 (−1.51 to −0.50) | −0.68 (−1.20 to −0.17) | −1.39 (−2.18 to −0.60) | 0.66 (0.15 to 1.16) |
|  |  | *q* = 0.417 | *q* = 0.222 | *q* = 0.881 | *q* = 0.305 | *q* = 0.881 |
| + hypertension | 101 | −0.83 (−1.36 to −0.29) | −0.88 (−1.40 to −0.36) | −0.33 (−0.86 to 0.19) | −1.23 (−1.96 to −0.49) | 0.94 (0.44 to 1.44) |
|  |  | *q* = 0.425 | *q* = 0.236 | *q* = 0.996 | *q* = 0.256 | *q* = 0.112 |
| + systolic blood pressure | 99 | −0.89 (−1.40 to −0.38) | −1.04 (−1.54 to −0.54) | −0.57 (−1.07 to −0.07) | −0.85 (−1.57 to −0.13) | 0.79 (0.31 to 1.27) |
|  |  | *q* = 0.182 | *q* = 0.056 | *q* = 0.821 | *q* = 0.741 | *q* = 0.289 |
| + BMI | 99 | −0.88 (−1.38 to −0.37) | −1.00 (−1.49 to −0.50) | −0.43 (−0.92 to 0.07) | −0.86 (−1.58 to −0.15) | 0.82 (0.35 to 1.30) |
|  |  | *q* = 0.186 | *q* = 0.060 | *q* = 0.991 | *q* = 0.779 | *q* = 0.183 |
| + pre-biopsy RAS inhibitor | 99 | −0.80 (−1.28 to −0.31) | −0.86 (−1.32 to −0.40) | −0.42 (−0.89 to 0.05) | −1.24 (−1.94 to −0.54) | 0.91 (0.47 to 1.36) |
|  |  | *q* = 0.539 | *q* = 0.157 | *q* = 0.971 | *q* = 0.298 | *q* = 0.086 |
| Two non-DKD participants with diabetes excluded | 119 | −0.95 (−1.43 to −0.47) | −1.09 (−1.54 to −0.63) | −0.66 (−1.12 to −0.19) | −1.19 (−1.96 to −0.41) | 0.57 (0.12 to 1.02) |
|  |  | *q* = 0.061 | *q* = 0.007 | *q* = 0.494 | *q* = 0.417 | *q* = 0.588 |

DKD minus the participant-frequency-weighted mean of other glomerular diseases, as log<sub>2</sub> fold change (95% CI). All estimates except the RAS inhibitor row include age, sex and batch; that row adds pre-biopsy renin–angiotensin system inhibitor use to the basic model (diagnosis only) in the 99 disease participants with medication records and expression data. The base estimates from the corresponding complete cases are given below. eGFR is per 10 mL/min/1.73 m<sup>2</sup>, systolic blood pressure (SBP) per 10 mmHg and body mass index (BMI) per kg/m<sup>2</sup>. For +eGFR, +hypertension, +systolic blood pressure and +BMI, the corresponding complete-case base estimates for *GLP1R*/*GRIA2*/*PRDM6*/*REN*/*P2RX1* were respectively −0.90/−1.08/−0.65/−1.13/0.57 (*n*=121); −0.88/−1.02/−0.47/−0.90/0.85 (*n*=101); and −0.86/−0.99/−0.43/−0.87/0.82 (*n*=99 for both SBP and BMI). Without the drug term, the basic model in the same 99 participants gave −0.81/−0.98/−0.51/−1.12/0.93. Whole-panel *q* values apply within each comparison and subset. Data file S1 retains complete-case intervals and *q* values.

**Supplementary Table S6.** KPMP source population and access.

| Item | KPMP single-nucleus atlas |
|---|---|
| Source | Kidney Precision Medicine Project single-nucleus RNA-sequencing atlas [S2] |
| Access route | KPMP data portal (https://atlas.kpmp.org/); public download, no approval required for public files |
| Reference-kidney definition | Reference tissue from living-donor, tumor-nephrectomy and deceased-donor sources; the reference type was retained for each sample |
| Comparison | DKD versus reference kidneys; vascular smooth muscle (subclass VSMC), participant-level pseudobulk |
| Participants | 15 with DKD and 22 reference participants, each with at least ten smooth muscle nuclei |
| Smooth muscle nuclei | 622 in DKD and 892 in reference kidneys; *GLP1R*-positive, 5 and 42 |
| Arteriolar hyalinosis sample | 62 participants with a hyalinosis grade and known sex: 35 with DKD and 27 with other diagnoses |

Gene-level estimates for the five altered mural genes are shown with their 95% CIs in Fig. 5a, and the 115-gene comparison with this study in fig. S4. Participant-level nucleus and positive-nucleus counts are given in the KPMP counts worksheet of data file S1.

